# Inferring person-to-person networks of *Plasmodium falciparum* transmission: is routine surveillance data up to the task?

**DOI:** 10.1101/2020.08.24.20180844

**Authors:** John H. Huber, Michelle S. Hsiang, Nomcebo Dlamini, Maxwell Murphy, Sibonakaliso Vilakati, Nomcebo Nhlabathi, Anita Lerch, Rasmus Nielsen, Nyasatu Ntshalintshali, Bryan Greenhouse, T. Alex Perkins

## Abstract

Inference of person-to-person transmission networks using surveillance data is increasingly used to estimate spatiotemporal patterns of pathogen transmission. Several data types can be used to inform transmission network inferences, yet the sensitivity of those inferences to different data types is not routinely evaluated. We evaluated the influence of different combinations of spatial, temporal, and travel-history data on transmission network inferences for *Plasmodium falciparum* malaria. We found that these data types have limited utility for inferring transmission networks and may overestimate transmission. Only when outbreaks were temporally focal or travel histories were accurate was the algorithm able to accurately estimate the reproduction number under control, *R*_*c*_. Applying this approach to data from Eswatini indicated that inferences of *R*_*c*_ and spatiotemporal patterns therein depend upon the choice of data types and assumptions about travel-history data. These results suggest that transmission network inferences made with routine malaria surveillance data should be interpreted with caution.

## Introduction

Concomitant with improved epidemiological surveillance, there is growing interest to leverage the collected data to infer transmission networks for a wide range of pathogens and to use those inferences to inform public health efforts. Past studies have incorporated temporal data^1^ and spatial data^2–5^ to estimate pairwise probabilities of transmission between individual cases and to use those estimates to infer time-varying and spatially varying reproduction numbers, respectively. More recently, methods have been developed to incorporate this type of detailed, individual-level epidemiological data^6–8^ to infer transmission networks for infectious diseases of humans, including severe acute respiratory syndrome^9^ and tuberculosis^10^, and of animals, such as rabies^11^ and foot-and-mouth disease^12^.

In addition to the diseases for which these methods have been applied to date, there is a growing need to apply similar methods to malaria in near-elimination settings. As incidence of malaria declines within a country, transmission becomes more heterogeneous in space and time^13^. Focal areas of high transmission, known as “hotspots,” pose a serious risk of fueling resurgence if left untargeted, potentially reversing decades of progress towards elimination^14^. To this end, granular estimates of when and where transmission occurs are needed, as spatially aggregated estimates may obscure important heterogeneities of practical relevance to control efforts^15^. In addition to characterizing details of local transmission, measurement of progress towards malaria elimination hinges on correct classification of cases as imported or locally acquired^16,17^, which is a byproduct of estimating transmission networks.

Previous work on malaria has made progress on the use of individual-level epidemiological data to infer transmission networks of *Plasmodium falciparum*, the parasite primarily responsible for human malaria in many regions of the world. Churcher *et al*.^18^ used temporal data to estimate the proportion of imported cases needed to confidently estimate the reproduction number under control, *R*_*c*_, below one and thereby provide evidence of controlled, non-endemic malaria transmission. Reiner *et al*.^6^ then built upon this work by incorporating spatial data and inferring an individual-level transmission network of *P. falciparum* in Eswatini. More recently, Routledge *et al*.^19,20^ used related approaches to infer transmission networks and *R*_*c*_ of *P. vivax* in El Salvador and China.

As the adoption of these methods increases, in particular for malaria, care should be taken to assess how the epidemiological setting and the inclusion or exclusion of certain data types might affect the accuracy of transmission network inferences, as well as resultant inferences about epidemiological quantities including *R*_*c*_ and spatiotemporal variation therein. A recent study by Campbell *et al*.^21^ noted that epidemiological data alone was generally insufficient to reconstruct transmission networks of other pathogens, ranging from *Mycobacterium tuberculosis* to SARS-CoV. Although *P. falciparum* malaria was not considered in that analysis, its long serial interval^22^ calls into the question the utility of epidemiological data for this purpose, though this has been largely unaddressed in past studies. Furthermore, past transmission network inferences for malaria have relied on various types of epidemiological data, ranging from the timing of symptom onset^18–20^ to more detailed spatiotemporal data^6^. Each study incorporated travel-history information into transmission network inferences and considered these data to be perfectly accurate, assuming that all cases that reported travel were imported. However, travel history may be an imperfect indicator of importation owing to errors in recall^17^ and the fact that travel to an area of ongoing transmission alone does not guarantee that an individual was infected there^17,23^. *P. falciparum* transmission network inferences are likely to be sensitive to the choice of data types^24^, and failure to evaluate the sensitivity of transmission network inferences to choices about data types and different assumptions about possible errors in travel-history data could lead to apparently confident, though ultimately incorrect, assessments of *P. falciparum* transmission risk in near-elimination settings.

Here, we present a Bayesian method for inferring transmission networks based on temporal, spatial, and travel-history data for individual malaria cases. We use it to characterize the sensitivity of transmission network inferences to the inclusion of different data types and to different assumptions about the accuracy of travel histories. Our method builds upon previous work by leveraging individual-level epidemiological data to obtain posterior estimates of transmission networks and model parameters in a way that can accommodate different assumptions about errors in travel histories. After establishing a proof-of-concept of our inference method on simple test cases, we applied our method to real-world surveillance data from Eswatini and additional simulated data sets to understand how the inclusion or exclusion of different data types and different assumptions about travel-history error affect our ability to infer transmission networks and estimate transmission metrics, namely *R*_*c*_.

## Results

To establish proof-of-concept, we first applied our inference method on three simple test cases and evaluated how well our inferences recovered the true transmission networks. We then applied our method to surveillance data collected in Eswatini during 2013-2017. Our focus was less on understanding malaria epidemiology in Eswatini and more on understanding how epidemiological conclusions change with the inclusion or exclusion of different data types and different assumptions about travel histories. These inference settings used: (1) spatial and temporal data while estimating the accuracy of the travel history (default setting); (2) spatial and temporal data while believing the travel history; (3) spatial and temporal data alone; (4) temporal data while estimating the accuracy of the travel history; and (5) temporal data while believing the travel history. To validate the inferences based on data from Eswatini, we simulated data generated using posterior parameter estimates obtained from the data from Eswatini and evaluated the ability of our inference method to recover the true transmission networks along with the underlying parameters on those simulated data. Finally, we performed a simulation sweep across different epidemiological settings to determine the range of conditions under which our inference method yielded reliable estimates of transmission. A full description of the analyses and additional results can be found in the Supplement.

### Application to Eswatini surveillance data

We applied our method to surveillance data collected in Eswatini during 2013-2017. Under the default inference setting, we estimated the diffusion coefficient *D*, which quantifies the spatial spread of transmission, to be 4.42 km^2^ day^-1^ (2.92 – 6.18 km^2^ day^-1^) (Fig 1A). This corresponded to a median inferred transmission distance of 13.0 km (0.0160 – 65.9 km), a median inferred serial interval of 47 days (−33 – 150 days) (Fig 2A & 2B), and median estimates of τ_s_, the probability that an imported case reported travel, of 0.61 (0.44 – 0.78) compared to the prior distribution mean of 0.80 and τ_l_, the probability that a locally acquired case reported travel, of 0.57 (0.53 – 0.61) compared to the prior distribution mean of 0.20 (Fig 1B & 1C). That the 95% credible interval for τ_s_ contained 0.50 indicated that our inference algorithm found limited use of travel-history data in discriminating between imported and locally acquired cases, because that implies that imported cases have equal probabilities of reporting or not reporting travel. The algorithm estimated the proportion of imported cases to be 0.052, corresponding to *R*_*c*_ = 0.95. Mapping risk of importation and local transmission across Eswatini under the default inference setting, we estimated consistently low risk of importation throughout the country and transmission hotspots in the northeastern part of Eswatini, close to the border with Mozambique (Fig 3A & 3B).

**Fig 1.**
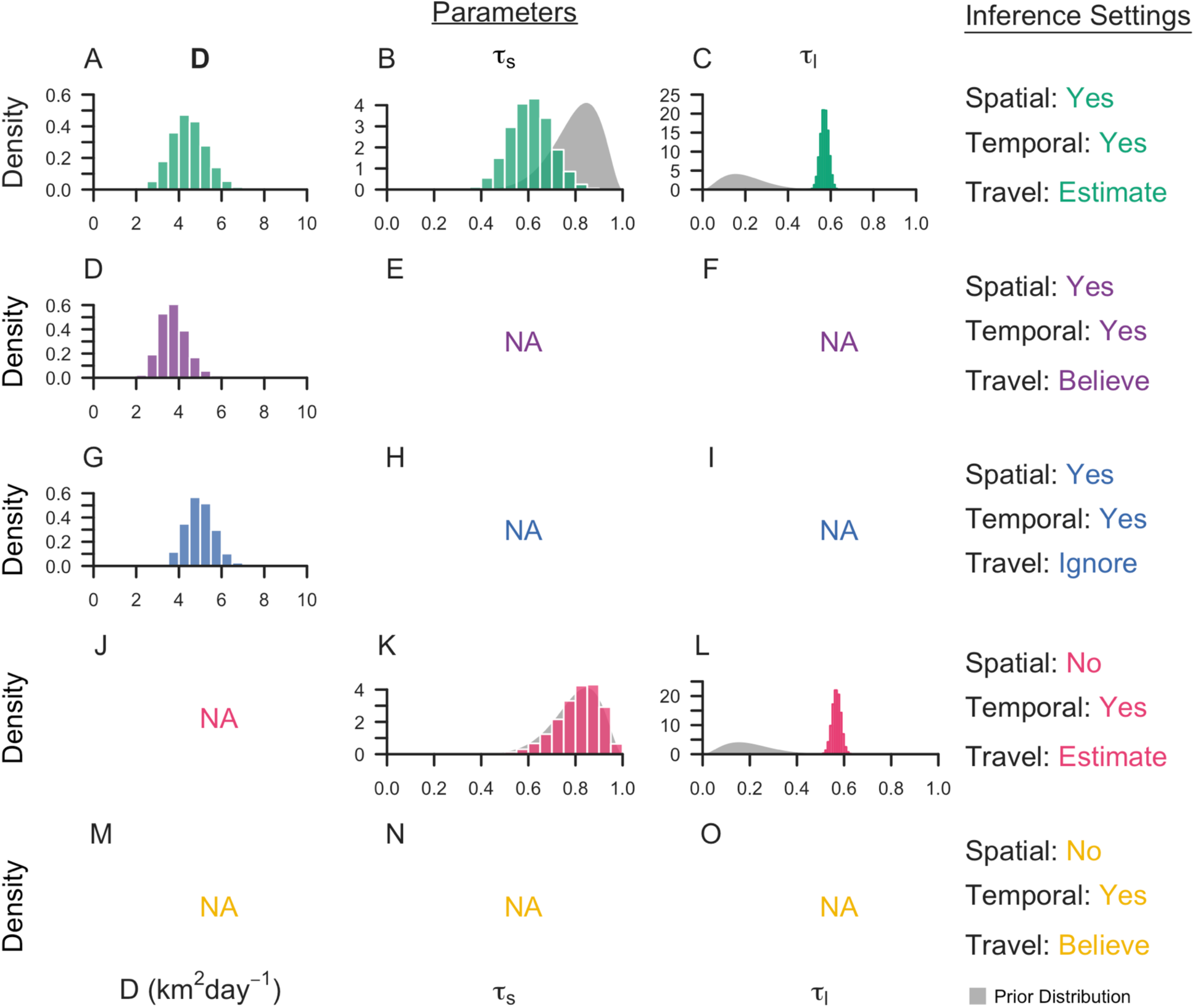
Marginal posterior distributions of parameters from Eswatini surveillance data. Histograms represent the marginal posterior distribution of each parameter, color-coded by the inference settings used. D is the diffusion coefficient with units km^2^day^-1^, τ_s_ is the probability that an imported case reports travel, and τ_l_ is the probability that a locally acquired case reports travel. Gray shapes represent the prior distributions placed on each parameter. Inference settings in which a given parameter was not estimated are indicated by NA.

**Fig 2.**
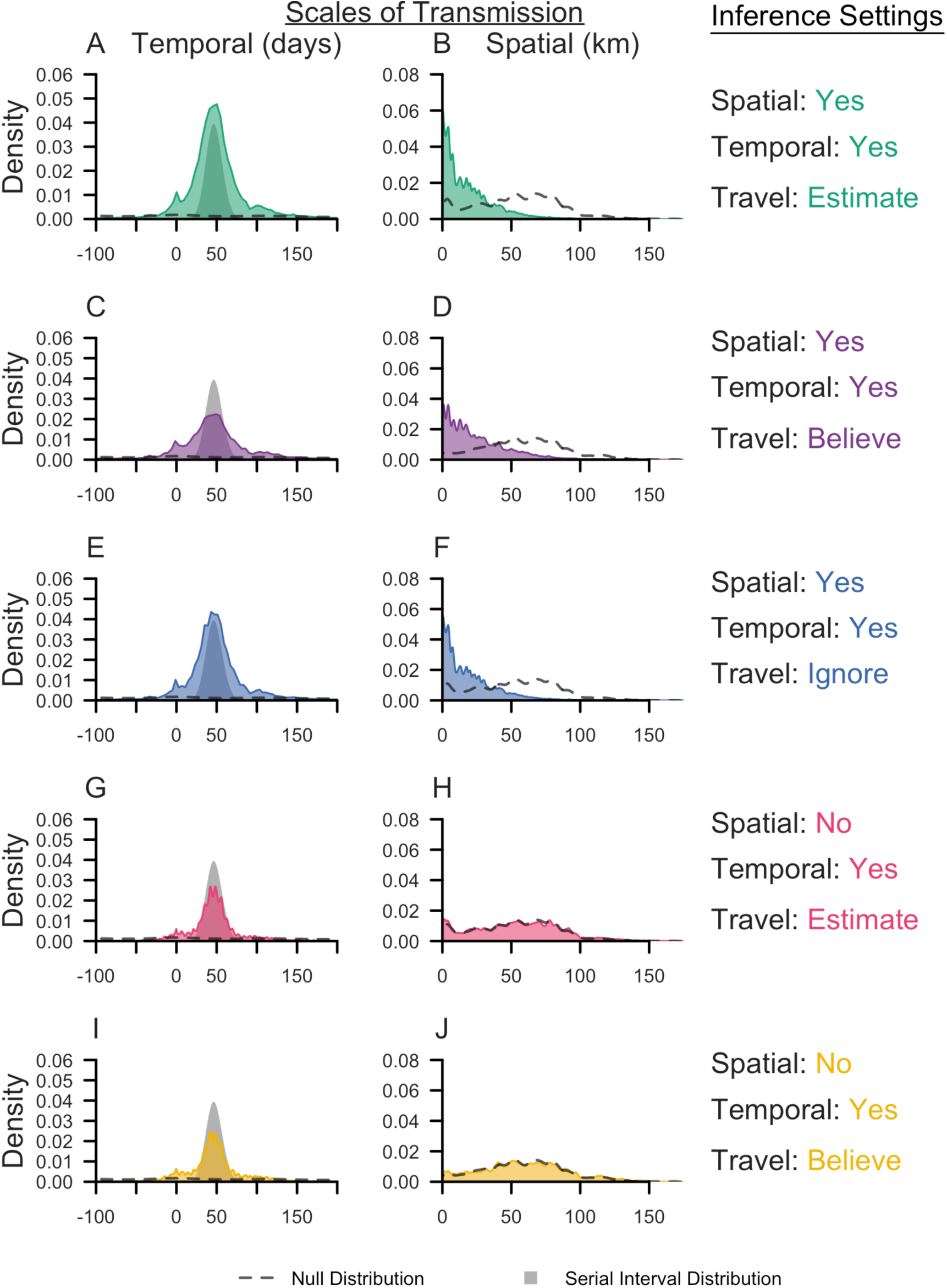
Spatial and temporal scales of transmission in Eswatini. Kernel density plots of the spatial (km) and temporal (days) scales of transmission are reported and color-coded for each inference setting. Dashed lines indicate the corresponding null distribution, generated from all random pairs of cases in the Eswatini surveillance data set. The null distribution was different if we believed the travel history, because classification of cases on the basis on travel history reduced the pairs of cases that could be randomly sampled. The grey shape is the serial interval distribution used in the likelihood.

**Fig 3.**
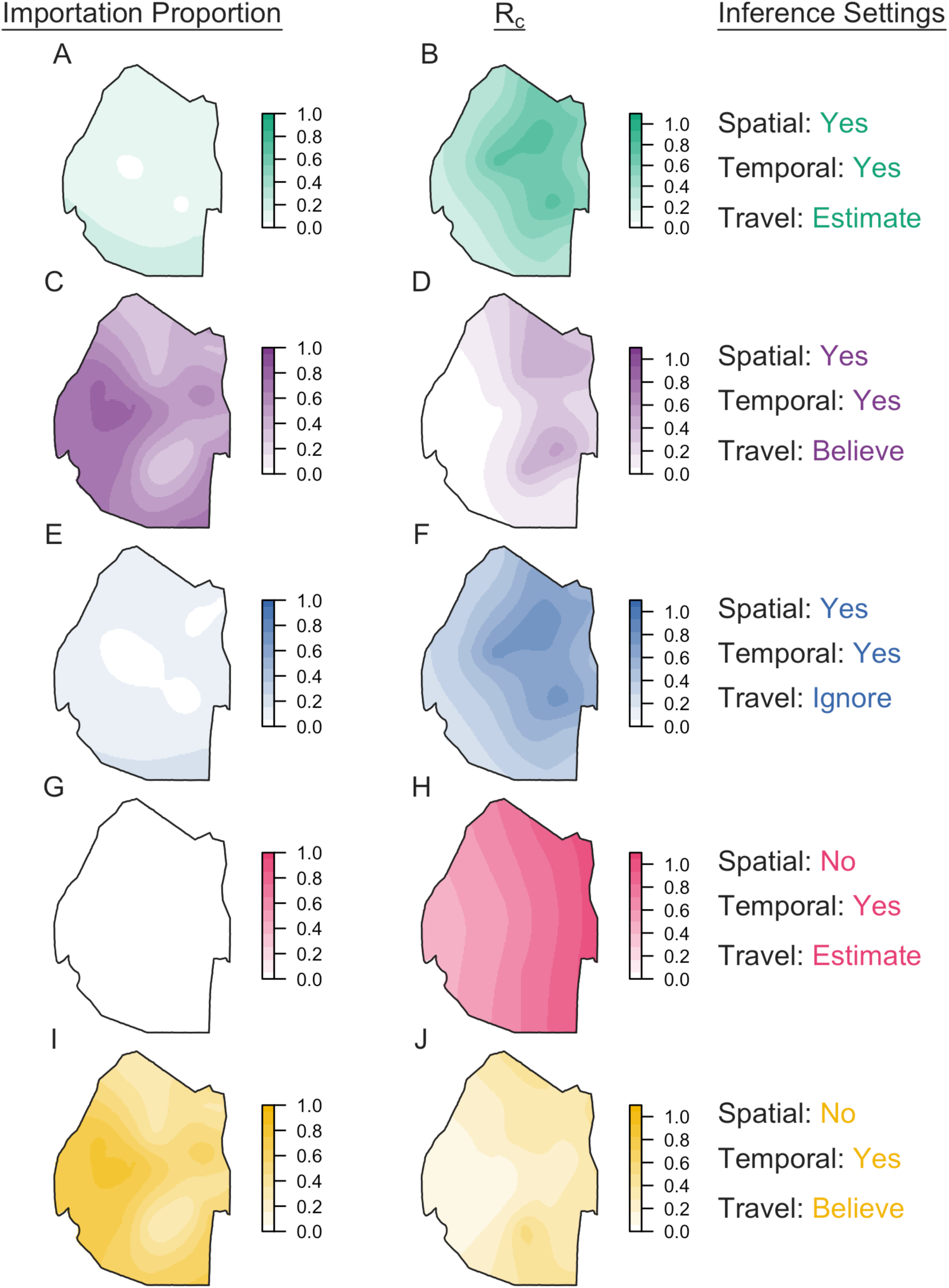
Spatial distribution of importation and transmission risk in Eswatini. Maps of the proportion of cases that are imported and the reproduction number under control (R_c_) were generated for each inference setting using a generalized additive model with a Gaussian process basis function setting using the mgcv package in R^25,26^. In each plot, darker colors indicate greater importation or transmission risk.

Parameter estimates and transmission network inferences differed under other inference settings. When we believed the travel history, we estimated a larger median transmission distance (Fig 2D). We attribute this increase in the spatial scale of transmission to clusters of cases with positive travel histories located near metropolitan areas. By forcing those cases to be imported, the algorithm tended to infer transmission across longer distances to explain the origins of the remainder of cases that did not report travel and were thereby inferred to be locally acquired. With respect to time, all five inference settings produced consistent serial interval estimates, though the inclusion of spatial data allowed for a wider range of transmission linkages in time (Fig 2A, 2C, & 2E). Finally, in the absence of spatial data, the model estimated higher predictive power of travel histories in identifying imported cases (τ_s_: 0.83, [0.60, 0.95]), though the travel history was consistently found to be uninformative for identifying locally acquired cases (τ_l_: 0.57, [0.53, 0.60]) (Fig 1K & 1L).

Classification of cases as imported or locally acquired, key information for control programs, was sensitive to the choice of inference setting. The proportion of cases classified as imported was most sensitive to different assumptions about the accuracy of the travel histories (Fig 3, left column; Fig 4). Believing the travel history yielded high estimates of importation in western Eswatini (Fig 3C & 3I), whereas estimating or ignoring the travel history yielded low, relatively homogeneous estimates of importation risk (Fig 3A, 3E, & 3G). For instance, using temporal data and estimating the accuracy of the travel history produced probabilities of importation that ranged 0.0043 – 0.0050, suggesting that nearly all cases resulted from local transmission (Figs 3G, 4D). Estimates of the spatial distribution of *R*_*c*_ depended most on the choice of which data types we included (Fig 3, right column). Notably, inclusion of spatial and temporal data produced a consistent spatial distribution of relative transmission risk, with transmission hotspots in northeastern Eswatini (Fig 3B, 3D, & 3F). However, believing the travel history reduced the magnitude of transmission that we inferred from a median *R*_*c*_ of 0.95 (Figs 3B, 4A) under default settings to 0.41 (Figs 3D, 4B). Omitting spatial data changed the spatial distribution of transmission. Estimating the accuracy of the travel history yielded high transmission estimates (median *R*_*c*_: 1.00) in eastern Eswatini (Fig 3H), whereas believing the travel history inferred hotspots of transmission (median *R*_*c*_: 0.42) in southern Eswatini (Fig 3J). We note that believing the travel history led to slightly different median estimates of *R*_*c*_ (0.41 vs. 0.42) depending upon whether spatial data was included, because the travel histories were unknown for 36 cases included in the analysis. As part of the inference procedure, the algorithm classified these cases as imported or locally acquired, and including spatial data caused a greater number of cases to be inferred to be imported.

**Fig 4.**
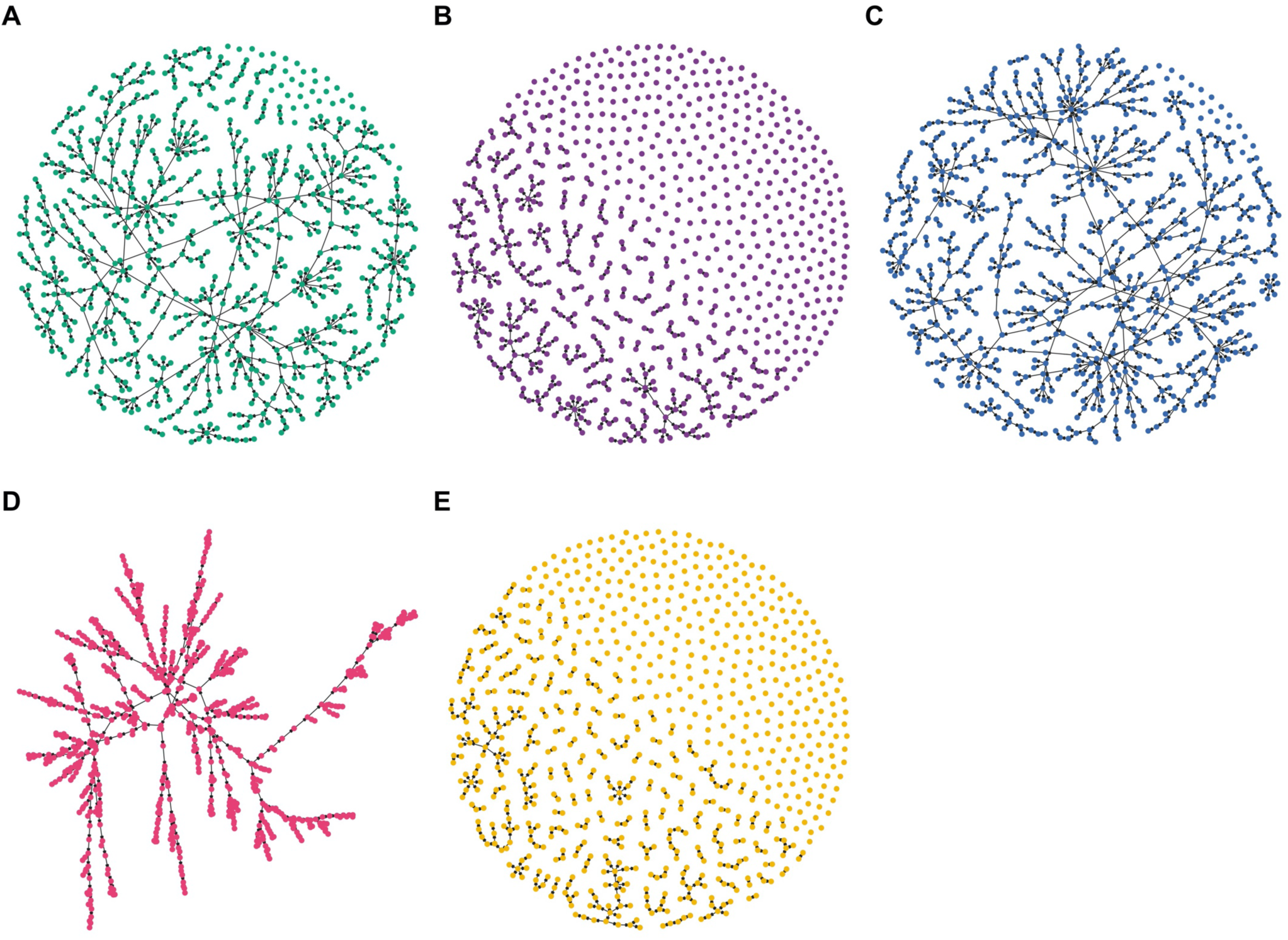
Maximum a posteriori transmission networks in Eswatini. The maximum a posteriori transmission networks (i.e., the transmission network in the posterior distribution with the highest likelihood) is shown for each inference setting: (A) spatial and temporal data while estimating the accuracy of the travel history; (B) spatial and temporal data while believing the travel history; (C) spatial and temporal data alone; (D) temporal data while estimating the accuracy of the travel history; and (E) temporal data while believing the travel history. In each transmission network, circles represent nodes, and arrows represent directed edges.

### Validation of inferences from Eswatini

Reconciling the different inferences under different inference settings in Figs. 1-4 was challenging because the true, underlying network and parameters were unknown. Using median posterior estimates from the Eswatini data under each inference setting, we simulated data and assessed the ability of the inference method to recover the underlying parameters and transmission networks (Table 1). We found that the model was able to estimate τ_s_ and τ_l_ reasonably well, depending on the inference setting (Fig 5). One exception was that the algorithm slightly overestimated τ_s_ under the default inference setting. We attribute this to the low proportion (0.052) of imported infections in the simulated data set and the strong prior placed on this parameter. This tendency was not observed under the inference setting where spatial data was excluded, because the true value of τ_s_ closely matched the mean of the prior distribution in that case (see the Supplement for further discussion).

**Table 1.**
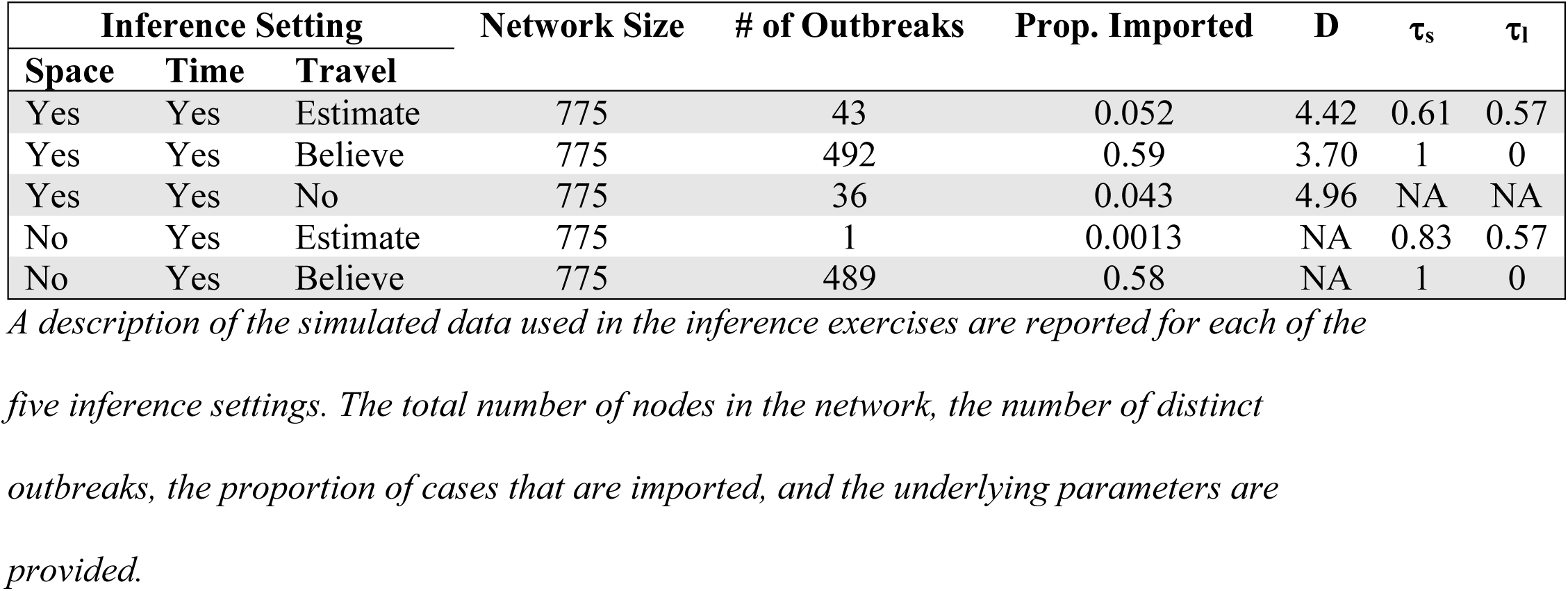
Characteristics of simulated data generated using the branching process model.

**Fig 5.**
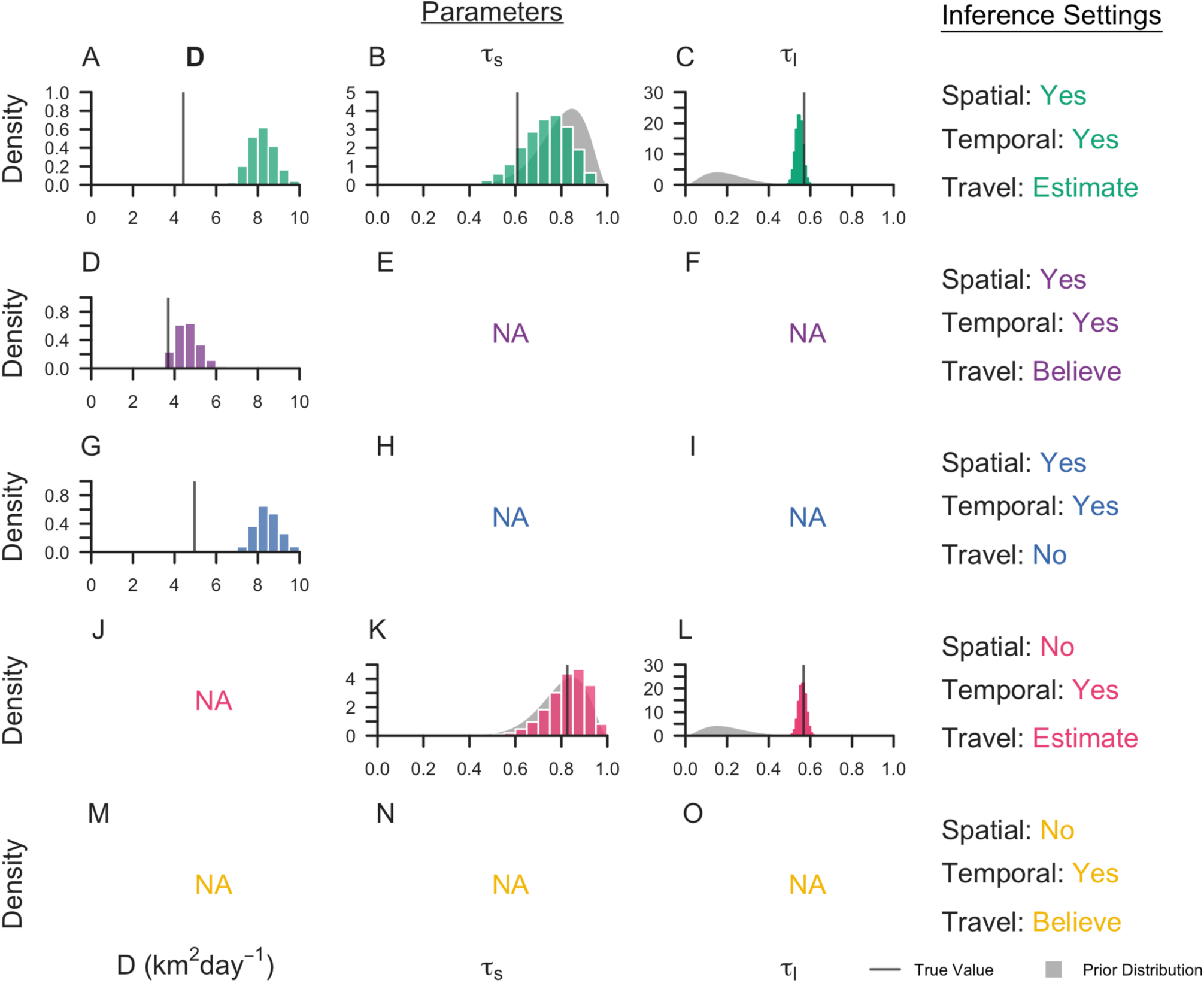
Marginal posterior distributions for parameters inferred from simulated data. The marginal posterior distributions are reported for each inference setting from its respective simulated data set. Each line denotes the true value of the parameter, and the grey shapes represent the prior distributions of the parameters. Inference settings in which a given parameter was not estimated are indicated by NA.

With the exception of believing the travel history, the model consistently overestimated the diffusion coefficient (Fig 5A, 5D, & 5G). We attribute the challenge of correctly estimating the diffusion coefficient to an inability to correctly estimate the underlying transmission network, the extent of local transmission in the network, and a numerical insensitivity in the overall likelihood to changes in *D*. When we conditioned the likelihood of *D* on the true transmission network when *R*_*c*_ was high, the true values of *D* fell close to the range of maximum-likelihood estimates, suggesting that this parameter could be estimated correctly if the true network was identified (S3 Fig). The likelihood around the true value was very flat, however, making it easy for *D* to be estimated incorrectly. When *R*_*c*_ was low, we underestimated the diffusion coefficient, because the likelihood of imported cases increases as *D* decreases.

The overall accuracy of classifying cases as imported or locally acquired was close to one (Fig 6). Though seemingly promising, these high accuracies masked a tendency to overclassify cases as locally acquired, because many more cases were simulated to be locally acquired than imported. For example, under the default inference setting, the accuracy of correctly classifying imported cases was 0.023 (0.023 – 0.067). Similarly, the accuracies of identifying the parent of each transmission linkage were poor, despite simulating under the assumptions of the model, with accuracies ranging from 0.042 (0.022 – 0.065) when using temporal data and believing the travel history to 0.31 (0.28 – 0.34) when incorporating spatial and temporal data and estimating the accuracy of the travel history (Fig 6, circle points). This suggests that, as the number of cases increases within a fixed space-time window, the information content of routinely collected epidemiological data decreases and the method becomes incapable of correctly estimating the transmission network. Nevertheless, under some settings, the method was able to capture higher-order summaries of the network, such as case classification and *R*_*c*_ (Fig 6, square and diamond points).

**Fig 6.**
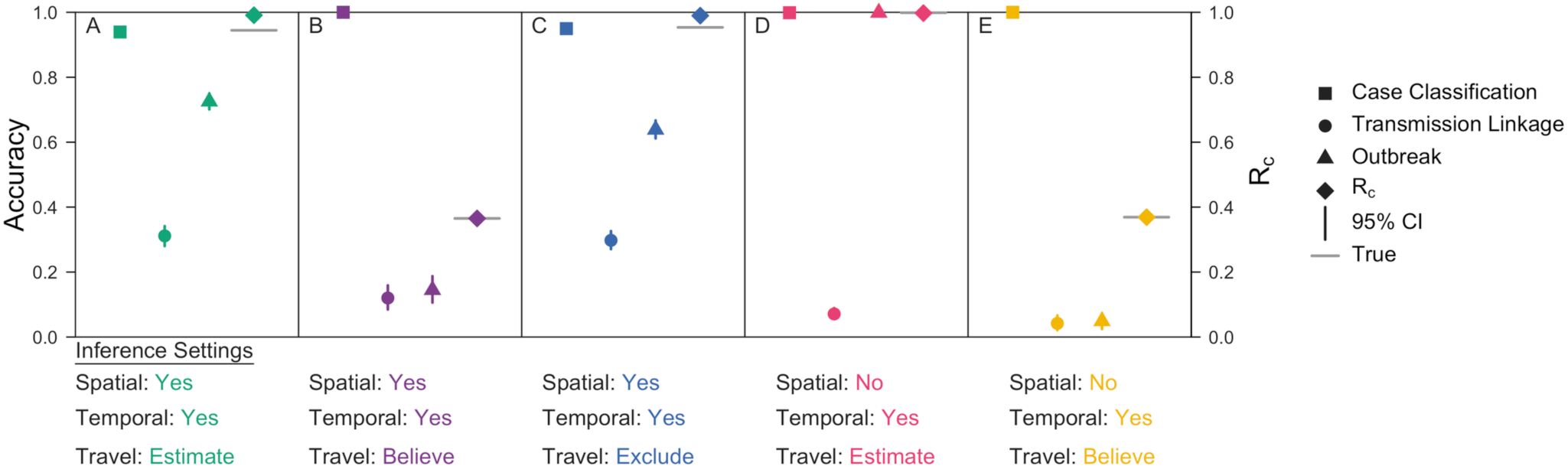
Inference accuracies for validation exercises. Accuracy metrics are reported for each inference setting applied to its respective simulated data set. Case Classification, represented by squares, refers to the proportion of cases that are correctly classified as imported or locally acquired. Transmission Linkage, denoted by circles, is the proportion of locally acquired cases for which the true parent is correctly identified. Outbreak, represented by triangles, is the proportion of locally acquired cases for which the inferred parent belongs to the correct outbreak. Bars denote the 95% credible intervals, and the grey line is the true R_c_ value of the network.

### Simulation Sweep

Validation of our inference algorithm revealed that its performance varied across simulated data sets. When applied to a series of simple test cases in which the transmission networks were small and in an optimal spatiotemporal arrangement, the inference method was able to reconstruct the transmission network and correctly estimate *R*_*c*_ (S2 Fig). When applied to larger transmission networks in which outbreaks overlapped in space and time, performance of the inference method was poor (Fig 6). This indicated that the performance of our inference algorithm depends on the epidemiological setting to which it is applied. To address this observation, we generated 2,000 simulated data sets in which we varied the proportion of imported cases, the spatiotemporal window over which imported cases were distributed, the diffusion coefficient, and the accuracies of the travel history (i.e., τ_s_ and τ_l_) (S2 Table). We then applied our inference algorithm under three different inference settings and quantified the accuracy of reconstructing each transmission network. The three inference settings used: (1) spatial and temporal data while estimating the accuracy of the travel history (default setting); (2) spatial and temporal data while believing the travel history; and (3) spatial and temporal data alone (S1 Table).

We observed that the accuracy of reconstructing transmission networks depended upon both the inference setting used and the epidemiological features of the simulated data. When we used spatial and temporal data and estimated the accuracy of the travel history or excluded it, the accuracy of reconstructing transmission networks depended on the relative proportion and temporal distribution of imported cases (S10 and S12 Figs). As the temporal window over which imported cases are distributed increased, the accuracy of identifying the true parent and the true outbreak of each locally acquired case increased. With an increasing temporal window, outbreaks within the transmission network became relatively more focal in time, which made the likelihoods of alternative transmission linkages more readily distinguishable. More accurate estimates of *R*_*c*_ under these inference settings similarly depended on the temporal window over which imported cases were distributed (Fig 7A, 7C). When the mean temporal interval between imported infections was greater than two times the mean length of the serial interval (i.e., approximately 100 days), our estimates of *R*_*c*_ improved, though we generally overestimated it. Furthermore, as the proportion of imported cases increased and *R*_*c*_ decreased, the accuracy of identifying the correct outbreak of each locally acquired case decreased (S10 and S12 Figs). This pattern reflected the relationship between *R*_*c*_ and the size of individual outbreaks. As *R*_*c*_ decreased, the size of individual outbreaks decreased, and, consequently, the probability that the inferred parent of a locally acquired case belonged to the same outbreak decreased.

**Fig 7.**
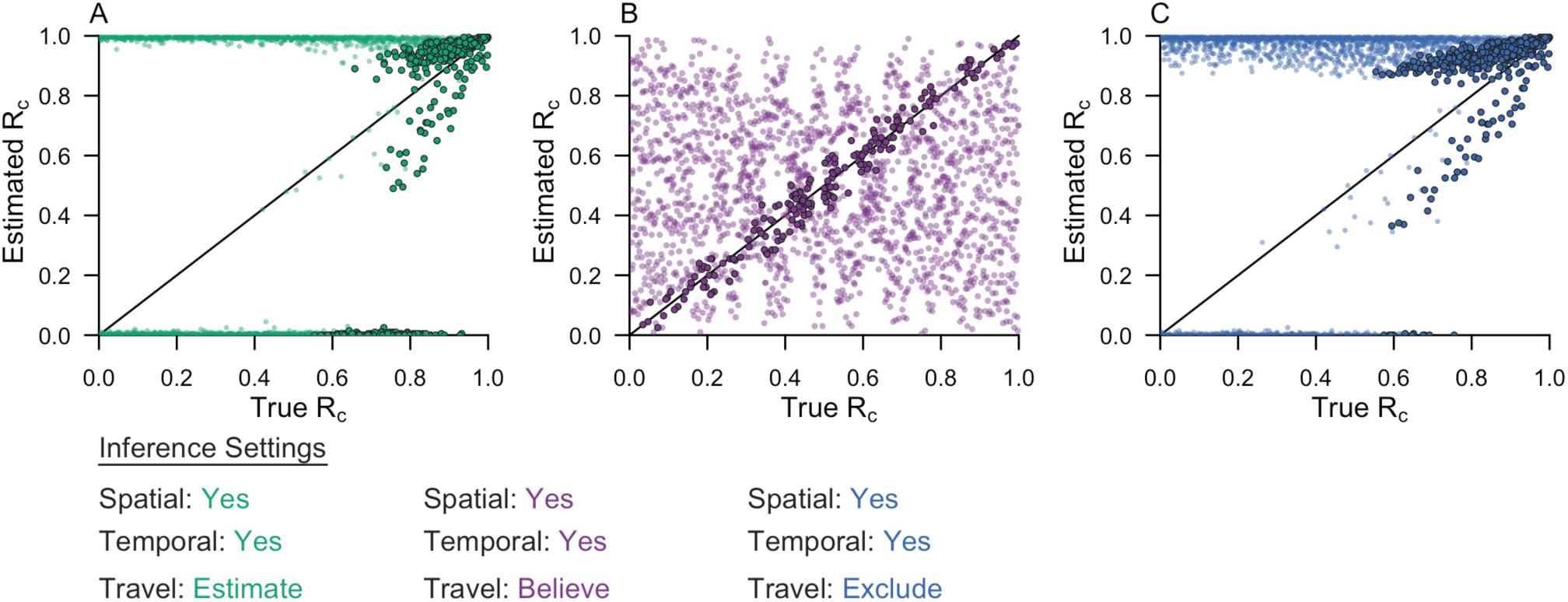
Comparison of R_c_ estimates across inference settings. The inference algorithm was applied to 2,000 simulated data sets. The estimated R_c_ is compared to the true R_c_ for each of the inference settings: (A) spatial and temporal data while estimating the accuracy of the travel history; (B) spatial and temporal data while believing the travel history; and (C) spatial and temporal data alone. Each point represents a simulated data set. The darker, accented points are simulated data sets with epidemiological features that improved performance. In (A) and (C), the darker, accented points were simulated data sets where the mean temporal interval between imported infections was greater than two times the mean serial interval. In (B), the darker, accented points were simulated data sets where the proportion of cases reporting travel was within 0.05 of the proportion of imported cases.

By contrast, when we believed the travel history, the accuracy of reconstructing transmission networks depended most strongly on the accuracies of the travel history. As the probability of reporting travel increased, the accuracy of classifying imported cases increased, and the accuracy of classifying locally acquired cases decreased (S11 Fig). Under this inference setting, our estimate of *R*_*c*_ depended only on the proportion of cases that reported travel. When the proportion of cases that reported travel matched the proportion of cases that were imported, we correctly estimated *R*_*c*_ (Fig 7B).

## Discussion

Our results show that, in most settings, routinely collected surveillance data offer limited value for reconstructing individual-level transmission networks of *P. falciparum* malaria and informing estimates of the reproduction number under control, *R*_*c*_. Using simulated data similar to the Eswatini surveillance data that we analyzed, we observed that our inference algorithm correctly identified transmission linkages less than 35% of the time. We attribute this inaccuracy primarily to the inherently limited information content of spatiotemporal data on *P. falciparum*. Its characteristically long serial interval^22^ means that an appreciable number of cases presenting within a short timeframe are difficult to link to each other based on their timing, even in a relatively facile test case in which the generative process assumed in the likelihood function matched that used to simulate the data. The inability to reconstruct transmission networks using routine surveillance data has been observed for other inference algorithms when applied to pathogens, such as *Mycobacterium tuberculosis* and *Klebsiella pneuomoniae*, with similarly long serial intervals, providing further evidence that the limitations noted in this study are generally inherent to the epidemiological data, rather than our method per se^21^.

Under most simulated scenarios and assumptions about the accuracy of travel-history data, we overestimated the number of locally acquired cases, leading to overestimates of *R*_*c*_. Crucially, our simulation sweep demonstrated that routinely collected surveillance data was most informative of individual-level transmission networks and *R*_*c*_ when local outbreaks were highly focal in time. Otherwise, while we were able to reconstruct the true transmission network with modest accuracy, we tended to misclassify truly imported cases as locally acquired, thereby overestimating *R*_*c*_. Taken together, these results suggest limited use of routinely collected surveillance data for informing fine-scale estimates of *P. falciparum* transmission. At broader spatial scales, however, routinely collected surveillance data may still have practical value, because the spatial distribution of cases can reveal epidemiological risk factors relevant for targeted control interventions^27,28^.

Although we were able to reach some general conclusions about our inference algorithm, our inferences were highly sensitive to which data types we included and which assumptions we made about the accuracy of travel-history data. Applying our algorithm to surveillance data from Eswatini, we observed that inferred patterns of transmission depended on which data types we included. With the inclusion of spatial data, we captured a spatial pattern of transmission consistent with another analysis from Eswatini^29^ with data from a different time period. Assumptions about the travel history appeared to have a strong influence on the overall magnitude of transmission that we inferred, due to the direct relationship between *R*_*c*_ and the proportion of imported cases^16^. As a result, believing the travel history, and thereby treating it as perfectly accurate as in previous approaches^6,18–20^, could bias *R*_*c*_ estimates if there are errors in travel-history data. A study comparing community travel surveys to mobile-phone data in Kenya found that travel histories considerably underestimated the volume of travel, suggesting high rates of false negatives in community travel surveys^30^. Believing the travel history may, then, underestimate the number of imported cases and overestimate *R*_*c*_. Accounting for inaccuracy in travel-history data is therefore important, and studies pairing community travel surveys with mobile-phone data could be used to inform prior distributions on the likely accuracy of travel-history data^30,31^.

The method that we used only considered a single spatial model to infer transmission linkages and assumed complete observation of cases, both of which are factors that could have affected our inferences based on the Eswatini surveillance data. The diffusion model that we used to represent spatial dispersion of parasites assumed that movement is isotropic in space and did not consider landscape features, such as heterogeneity in human population densities and environmental factors that may affect mosquito ecology. A study analyzing self-reported movement patterns in Mali, Burkina Faso, Zambia, and Tanzania found that gravity and radiation models of spatial dispersion fit the data well, though the appropriateness of each model depended on the type of traveler, the travel distance, and the population size of the destination considered^32^. Although a variety of spatial kernels could have been used in our analysis, we expect that the conclusions that we reached are robust to the choice of spatial kernel, because the spatial kernel used in the likelihood matched that used to simulate the data. Regarding the representation of *P. falciparum* infections in our data set from Eswatini, there are asymptomatic and mild infections that are unlikely to have been recorded in the surveillance system yet may comprise a substantial proportion of malaria infections within Eswatini ^13^. Accordingly, it is possible that our assumption of complete observation of cases could have biased *R*_*c*_ estimates, likely downward due to the fact that missing cases will tend to make offspring numbers appear smaller than they actually are^33,34^. Even so, we expect that our conclusions about the sensitivity of transmission network inferences to the choice of data types and assumptions about travel-history data are robust to these limitations of our study. This further reinforces our conclusion of the need for caution in attempting to reconstruct person-to-person transmission networks from routine surveillance data^35^, because incomplete observation of cases would lead to greater inaccuracies in our transmission network inferences beyond what we noted in our study.

Given that some of the limitations of our approach may be inherent to the information content of these data types in this system, one potential avenue for improving inferences of fine-scale patterns of *P. falciparum* transmission could involve the integration of additional data streams. For example, mobile-phone data^36^ and high-resolution friction surfaces^37^ could more realistically characterize mobility patterns, whereas travel-history information that details the dates, duration, and location of each trip that has been used in programmatic contexts^27^ could more accurately identify importation events. Additionally, the inclusion of pathogen genetic data, which has the potential to provide a more direct signal of parasite movement, could complement traditional epidemiological data^38^. Diverse genetic markers of *P. falciparum* have been characterized in near-elimination settings, such as Eswatini^39^, and have been successfully used to identify imported cases in Bangladesh^40^ and Namibia^31^. There is also scope for further methodological development, such as relaxing our assumption of complete observation of infections and incorporating an underlying mechanistic model of transmission (as in Lau *et al*.^8^; Guzzetta *et al*.^41^). Incorporating an underlying mechanistic model would relax our uninformative prior assumption on all possible transmission networks, ruling out transmission networks that are epidemiologically implausible and allowing us to account for spatial differences in transmission potential and the rate of importation due to different epidemiological and demographic factors. This approach would also permit us to estimate the serial interval distribution and seasonal variation therein directly from the data rather than borrow estimates from the literature^22,42,43^. To this end, we envision that leveraging the strengths of our method along with other, complementary methods could strengthen inferences based on routinely collected epidemiological data and open up new possibilities to make use of even more data types, such as serological data, prevalence surveys, and pathogen genetic data^38,44,45^.

## Conclusions

We demonstrated the limitations of routinely collected surveillance data for the inference of individual-level transmission networks of *P. falciparum*. We identified a tendency to overestimate local transmission using routinely collected surveillance data, especially when outbreaks overlapped in space and time. Using both real data from Eswatini and simulated data, we identified strong sensitivities of our inferences to the epidemiological setting, the choice of data types included, and assumptions about the accuracy of travel-history data. Our results indicated that using spatial and temporal data and believing travel histories yielded the most plausible estimates of transmission when applied to the Eswatini surveillance data. However, our simulation sweep demonstrated that the accuracy of our inferences strongly depended on the accuracy of the travel-history data when the travel-history data were assumed to be accurate. These sensitivities to the choice of data types and assumptions about the accuracy of travel-history data could have important programmatic implications if outputs of transmission network inferences are operationalized. Although this study was specific to *P. falciparum*, the results of our analyses indicate that future studies inferring transmission networks of *P. falciparum*, or any pathogen, should carefully consider the epidemiological setting and the choice of data types and assumptions that inform the model and should validate them using simulated data.

## Methods

### Bayesian framework for estimating transmission linkages

Our goal was to obtain probabilistic estimates of a transmission network *N* that defines transmission linkages among a set of known cases. The transmission network is defined as a directed, acyclic graph comprised of a set of directed edges represented as *N =* {*N*_*i,j*_} for all *i, j*. Each *N*_*i,j*_ indicates that case *i* is hypothesized to contain parasites that are the most direct observed ancestors of the parasites contained in case *j*. In addition, at least one edge denoted *N*_*u,j*_ must exist in the network, indicating that the parasites contained in case *j* have no ancestors among the parasites contained in any known local case and are instead contained in some unknown case *u* from some source population *s*, such that it is denoted *u*_*s*_. To illustrate this terminology, an example transmission network is depicted in Fig 8.

**Fig 8.**
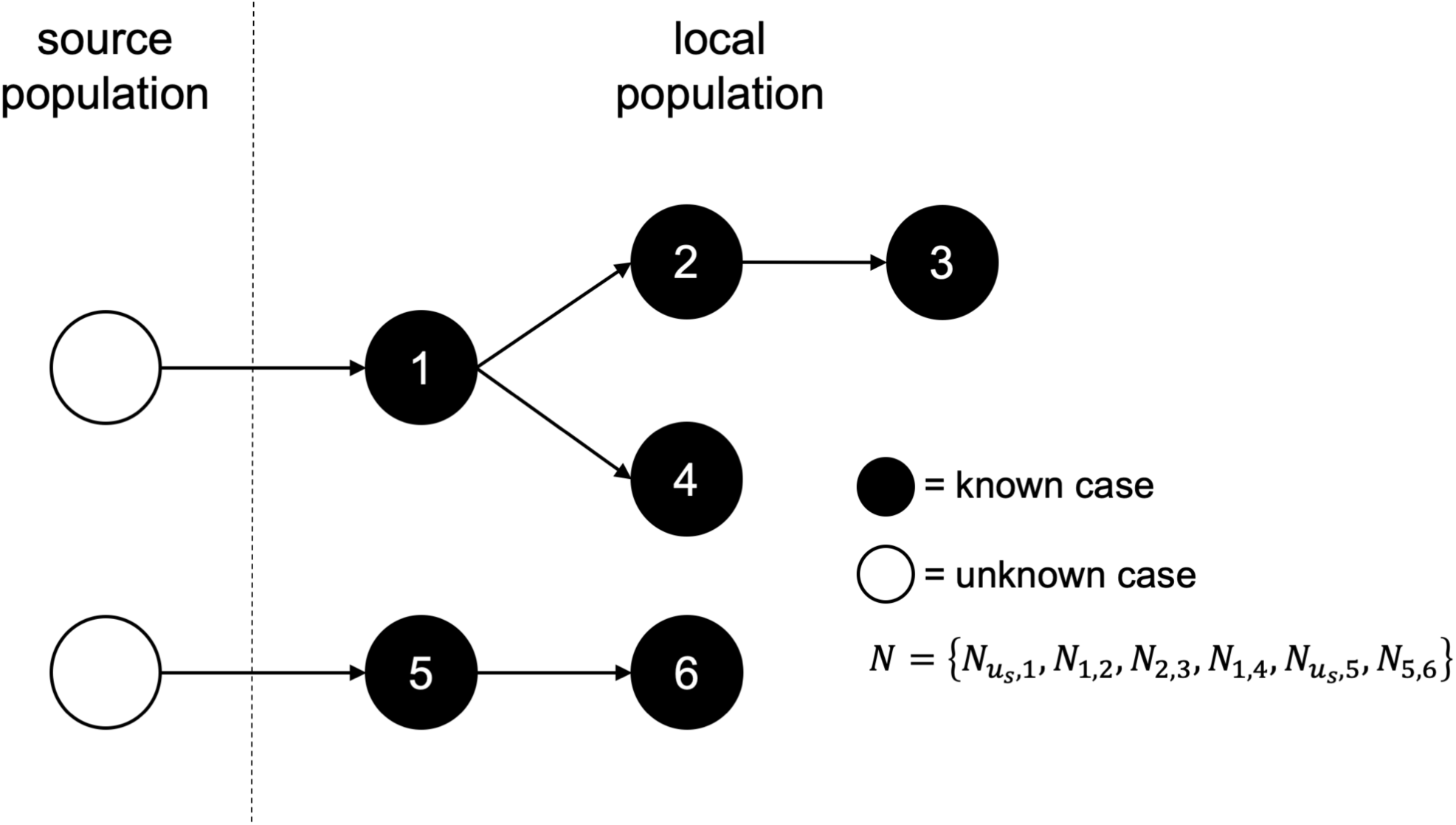
Schematic of a hypothetical transmission network. A hypothetical transmission network is presented along with the corresponding notation. In the schematic, white circles denote unobserved cases, and black circle denote observed cases. Arrows represent transmission between two cases.

To estimate *N*, we used spatial, temporal, and travel-history data about all cases, denoted as 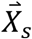, *X*_*t*_, and *X*_*h*_, respectively. We did so within a Bayesian statistical framework, meaning that we sought to estimate the joint posterior probability density,

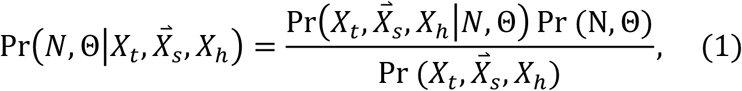

of the transmission network defined by *N* and the model parameters Θ conditional on the data 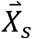, *X*_*t*_, and *X*_*h*_. The first term in the numerator of eq. (1) is the likelihood of *N* and Θ conditional on the data. The second term in the numerator is the prior probability of *N* and Θ. The term in the denominator is the probability of the data, which is an intractable quantity to calculate directly given that it would require evaluation of an extremely high-dimensional integral over *N* and Θ. To address this, we used a Markov chain Monte Carlo algorithm to draw random samples of *N* and Θ from the posterior distribution specified in eq. (1).

The most critical piece of our inference framework is the likelihood, which we define as a function of each case *j* as

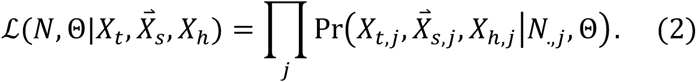

Below, we define the probability of the data associated with each known case *j* as a function of different assumptions that are possible about how case *j* is connected to the rest of the transmission network.

### Scenario 1: Local transmission between known cases *i* and *j*

When case *i* contains parasites that are immediate ancestors of the parasites contained in case *j*, we represent its contribution to the likelihood as

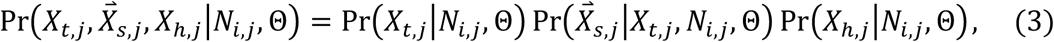

which is the product of the probabilities of the temporal, spatial, and travel-history data given the network and model parameters. This formulation assumes that those data are generated independently for each individual, with the exception of a dependence of the spatial data on the temporal data.

#### Probability of the temporal data

To characterize the time elapsed between two cases resulting from local transmission, we used a model of the generation and serial intervals for *P. falciparum* malaria by Huber *et al*.^22^. The generation interval represents the time between infection of a primary and secondary case, whereas the serial interval represents the time between detection of those cases. Because the timing of infection per se (i.e., an infectious mosquito inoculating a susceptible human) is typically unknown, we focused on the serial interval as the most apropos temporal quantity relating cases.

In deriving the probability of a given length of the serial interval, Huber *et al*.^22^ convolved a random variable representing variability in the generation interval (*GI*) with a random variable representing variability in the time between infection with *P. falciparum* and detection by surveillance – i.e., the infection to detection period (*IDP*). That framework yielded

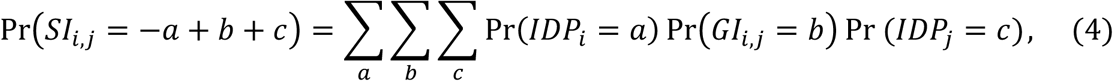

as the probability of a serial interval of length *SI*_*i,j*_. We allowed for different models of the serial interval depending upon differences in the *GI* and *IDP* for different types of primary and secondary cases. For instance, we assumed that symptomatic cases present in a clinic some number of days after infection as informed by empirical data from Zanzibar^22^. For an asymptomatic infection, we assumed that detection occurred through active surveillance at a randomly drawn day among all days where its asexual parasitemia exceeds a detection threshold^22^. The choice of *IDP* for both the primary and secondary case informs the probability of two cases separated in time by *SI*_*i,j*_ = *X*_*t,j*_ − *X*_*t,i*_ days.

#### Probability of the spatial data

Following Reiner *et al*.^6^, we assumed that a simple two-dimensional Wiener diffusion process determines the location of secondary cases relative to the location of their associated primary case. It follows that, for a given diffusion coefficient *D* with units *km*^*2*^*day*^*-1*^ and generation interval *GI*_*i,j*_, the two-dimensional location 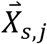 of the secondary case *j* is described by a bivariate normal distribution with probability density

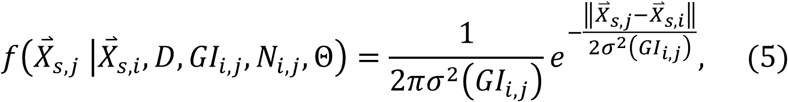

where *σ*^2^(*GI*_*i,j*_) *= DGI*_*i,j*_. This formulation assumes that each spatial dimension is independent, that the variance scales linearly with the generation interval, and that movement is isotropic across a continuous landscape.

One complication to eq. (5) is that the generation interval *GI*_*i,j*_ is unobserved and, therefore, cannot take on a fixed value. Instead, we must use data about the serial interval *SI*_*i,j*_ to inform our generative model for 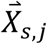. To do so, we take advantage of the property of normal random variables that the sum of two or more random variables is itself a normal random variable^46^. This property allows us to recast eq. (5) as a function of *SI* rather than *GI* by computing the appropriate σ^2^ as

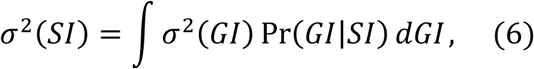

which is effectively a weighted sum of the spatial variances associated with a given *GI* proportional to the probability of each *GI* conditional on the observed *SI*. This results in

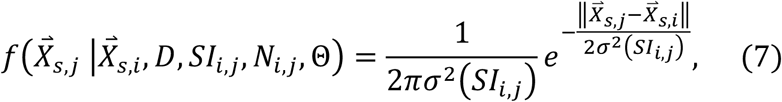

as the probability density of the spatial data that we assume.

In the event that case *i* has missing spatial data, we cannot compute the spatial likelihood of eq. (7). To address this, we define a latent unobserved quantity 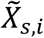, which represents the unknown location of case *i*. We then integrate over the uncertainty in 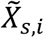,

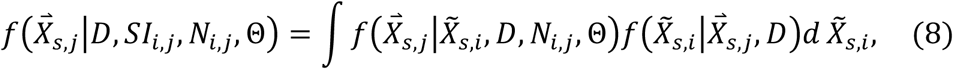

to compute the probability density of case *j* with known spatial location 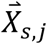 arising from case *i* with unknown spatial location 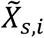. Equation (8) is computed as the product of the probability density of the location of a known case *j* conditional on an unknown location 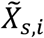 and the probability density of spatial separation 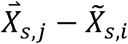 conditional on the diffusion coefficient *D* for all 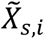 Because we assume that movement is isotropic, eq. (8) is a two-dimensional Gaussian integral, simplifying to

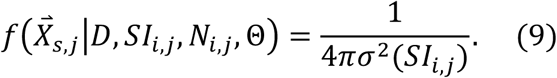

In the event that case *j* has missing spatial data and case *i* has known spatial data, the latent unobserved quantity becomes 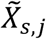. We then integrate over the uncertainty in 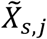 and calculate 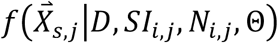 using eq. (8-9).

#### Probability of the travel-history data

Although we assume in this scenario that a person’s infection was locally acquired, our model must still be capable of explaining the travel-history data *X*_*h,j*_. We define a probability τ_l_ that case *j* reported travel (i.e., *X*_*h,j*_ = 1) even though they were not infected during that period of travel, such that

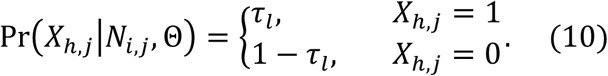

In the event that case *j* has missing travel-history data, we cannot compute the travel-history likelihood of eq. (10). To address this, we defined a latent unobserved quantity 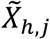, which represents the unknown travel history of case *j*. We then sum across the uncertainty in 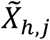,

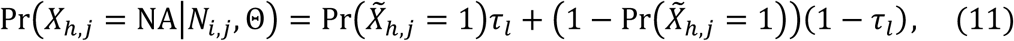

to compute the probability that case *j* was locally acquired given an unknown travel history. In eq. (11), 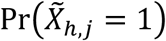 was computed as the proportion of cases with a positive travel history among all cases with known travel-history data.

Taken together with the probabilities of the temporal and spatial data described above, the product of these three probabilities constitutes the entirety of the contribution of a case *j* infected by a known local case *i* to the overall likelihood of *N* and Θ.

### Scenario 2: Importation of local case *j* from source population *s*

In the event of 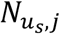, we represent the contribution of such a case to the overall likelihood of *N* and Θ as the product of the probabilities of its temporal, spatial, and travel-history data under similar assumptions as in Scenario 1. The key difference in this scenario is that there is no information about the unknown source case that gave rise to case *j*.

#### Probability of the temporal data

Because the person containing parasites that are the direct ancestors of those in case *j* is unobserved and does not have an *X*_*t,i*_, we are unable to compute the probability of the temporal data as described in Scenario 1. It is important though to obtain a probability comparable to that from Scenario 1 as a reference point for determining whether it is more likely that a given case arose from some other known local case or from an unknown case *u*_s_ from source population *s*. To do so, we consider the variable 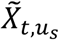, which is a latent variable describing the timing of when *u*_s_ would have been detected, had it been detected.

Because *u*_s_ is not observed, we considered it to be asymptomatic and untreated. We then calculated the probability of the timing of a known case *j* arising from an unknown case *u*_s_ as

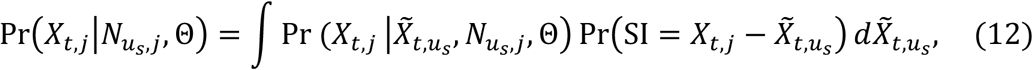

by integrating over uncertainty in 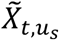. We represented this as the product of the probability of the timing of a known case *j* conditional on an unknown time of detection 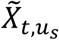 and the probability of the serial interval 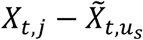 for all 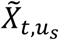. In equation (12), we did not distinguish between symptomatic and asymptomatic cases *j* because the calculation is identical; only the serial interval distributions differ.

#### Probability of the spatial data

Without an 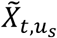 for the unobserved case *u*_s_, we lacked information on the serial interval between it and case *j*. Consequently, we were unable to use the probability from eq. (7) in that particular form. Instead, we computed the spatial variance as a function of the diffusion coefficient alone, yielding

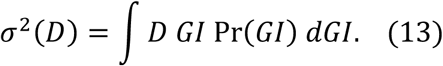

Equation (13) integrates across all possible generation intervals and simplifies to *D*𝔼[*GI*], the product of the diffusion coefficient and the expectation of the generation interval.

We applied this spatial variance to the unobserved latent variable 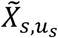, which represents the unknown location of the unobserved case *u*_s_. We integrated over uncertainty in 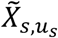 to compute the probability density,

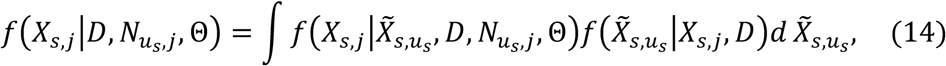

of the location of a known case *j* arising from an unknown source case *u*_s_ with unknown location 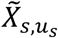. This is represented as the product of the probability density of the location of a known case *j* conditional on an unknown location 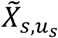 and the probability density of spatial separation 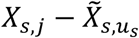 conditional on the diffusion coefficient *D* for all 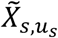. As in eq. (9), we treated eq. (14) as an evaluation of the Gaussian integral, evaluating to

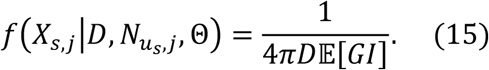

In eq. (15), *D* is the diffusion coefficient and 𝔼[*GI*] is the expectation of the generation interval.

#### Probability of the travel-history data

We considered the travel history *X*_*h,j*_ to be a binary variable with a value of 1 indicating reported travel to an area with known or assumed malaria transmission within a timeframe consistent with the person having become infected there. After defining the probability τ_s_ that *X*_*h,j*_ = 1 conditional on 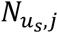 it follows that

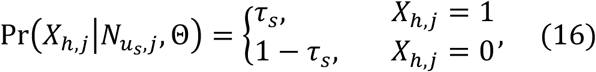

which constitutes the contribution of the travel history of such a case to the overall likelihood of *N* and Θ. If the travel history of case *j* is unknown, an analogous calculation to eq. (11) is made using τ_s_.

### Bayesian inference

#### Markov Chain Monte Carlo Algorithm

To avoid evaluating the high-dimensional integral over *N* and Θ, we drew samples of *N* and Θ from their posterior distribution defined by eq. (1) using a Metropolis-Hastings Markov chain Monte Carlo (MCMC) method^47,48^. To begin the chain, *N* and Θ were initialized to *N*^(1)^ and Θ^(1)^, and each subsequent step *i* in the chain was denoted *N*^(*i*)^ and Θ^(*i*)^. At each step, states *N*^′^ and Θ^′^ were proposed with Pr ((*N*^(*i*)^, *Θ*^(*i*)^)*→* (*N*′, *Θ*′)). Proposed states were accepted with probability

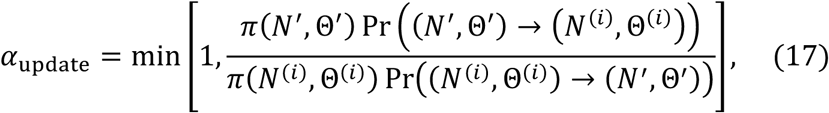

where π(*N, Θ*) is the product of the likelihood 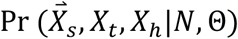 of *N* and Θ conditional on the data and the assumed prior probability Pr (*N, Θ*) of *N* and Θ. After a random draw *R* from a uniform distribution, the chain was updated according to

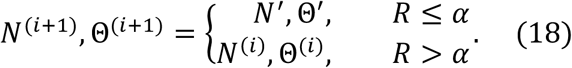

To reduce the probability of the chain becoming stuck at a local maximum, we employed Metropolis-coupled Markov chain Monte Carlo (MC^3^)^49^. Implementing MC^3^ involved running multiple chains in parallel, with π_c_(*N, Θ*) in chain *c* raised to the power β_c_ according to

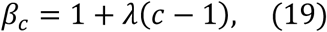

where λ > 0 is a temperature increment parameter that governs the degree to which each chain is “heated.” As a result of setting β_1_ = 1, π_1_(*N, Θ*) is directly proportional to the joint posterior distribution and is referred to as the master or “cold” chain. This algorithm effectively flattens the likelihood in the heated chains by setting *β*_c_ > 1, allowing them to explore the parameter space more freely and to encounter alternative high-density regions more readily than the cold chain would alone. At a pre-defined frequency, two randomly selected chains *i* and *j* were allowed to swap parameter sets according to a swap probability

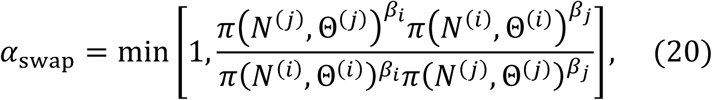

where π(*N, Θ*) is the same as it was in eq. (17). A swap into the master chain only occurred if it was from one of the two randomly selected chains and *R* ≤ *α*_swap_. We recorded a total of 100 million samples from the posterior distribution, discarding the first 50 million samples as burn-in and thinning the chain every 10,000 samples between each recorded sample.

#### Proposals

Proposals made by the MC^3^ algorithm involved changes to the parameters (i.e., *D*, τ_s_, and τ_l_) and changes to the transmission network topology. Each proposal occurred with a fixed probability, where the sum of these proposal probabilities was equal to one.

Proposals to change parameters involved updating *D*, τ_s_, or τ_l_. To update the value of *D*, a new value was drawn from a normal distribution with mean set to the current value of the parameter and variance set to 2.5. Values of *D* proposed must be strictly nonnegative, so we rejected any proposed *D* that was less than zero and assigned *α*_update_ *=* 0. Similarly, new values of τ_s_ and τ_l_ were chosen according to normal distributions with means set to their current parameter value and variance set to 0.25. Because τ_s_ and τ_l_ are probabilities, we rejected any proposed value that fell outside the range [0,1] and assigned *α*_update_= 0.

Changes proposed to the network topology involved the addition or removal of an ancestor from a randomly selected node. We assigned a uniform probability of proposing case *a* as an ancestor to a randomly selected case *i*, such that proposals to the network topology are uninformed by spatial and temporal data. Furthermore, we defined the proposal probability of removing case *a* as an ancestor to a randomly selected case *i* as

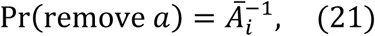

where *Ā*_*i*_ represents the size of set *A*_i_ of all ancestors to case *i*. Proposed changes to the network are then accepted according to eq. (17).

#### Prior assumptions

We placed strong priors on τ_s_ and τ_l_, because we assumed that travel histories were mostly, but not completely, accurate. We used a beta-distributed prior on τ_s_, with parameters 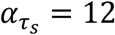 and 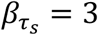, which resulted in a mean of 0.8 and a variance of 0.01 for this prior distribution. We also used a beta distributed prior on τ_l_, with parameters 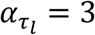 and 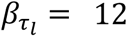, which resulted in a mean of 0.2 and a variance of 0.01. We assumed a uniform prior on *D* over the interval [10^−3^, ∞) and an even prior across all possible network configurations, meaning that those prior probabilities canceled out in eqs. (17) and (20).

#### Assessing convergence

For *D*, τ_s_, and τ_l_, we assessed convergence using the Gelman-Rubin statistic^50^, with values below 1.1 indicating convergence. For the transmission network *N*, we assessed convergence by calculating correlation coefficients of case-level probabilities across five chains from independent realizations of the MC^3^ algorithm, for a total of 10 pairwise comparisons across the five chains. The two case-level probabilities that we considered were the posterior probability that each case was infected by an unknown case *u*_*s*_ from a source population and the posterior probability that each case *j* was infected by each other case *i*. Higher values of these correlation coefficients provided stronger support for convergence.

#### Ethical Considerations

Ethical approval was obtained from the Eswatini Ministry of Health, the University of California, San Francisco, and the University of Notre Dame (IRB 19-06-5408). All data were analyzed anonymously.

## Supporting information

Supplemental Methods and Results

## Data Availability

The code and simulated data to reproduce the analyses can be found at https://github.com/johnhhuber/SpaceTime_Networks. The data collected from Eswatini contains sensitive household locations and are unable to be shared due to institutional review board restrictions.

## Competing Interests

The authors have no competing interests to declare.

## Acknowledgments

The authors thank the National Malaria Elimination Programme as well as Nontokozo Mngadi and Deepa Pindolia from the Clinton Health Access Initiative for their support in the collection of the surveillance data used in this study. We also thank Brooke Whittemore for her support in data management. JHH acknowledges support from a National Science Foundation Graduate Research Fellowship and a Richard and Peggy Notebaert Premier Fellowship. BG and TAP received support from a grant from the Bill and Melinda Gates Foundation (OPP 1132226 to BG). MSH received support from NIAID (AI101012). The funders had no role in study design, analysis, decision to publish, or preparation of the manuscript.

## Author Contributions

J.H.H, T.A.P, and B.G. conceived of the study. M.S.H., N.D., S.V., N.N., N.N, and B.G. curated the data. J.H.H., M.S.H., M.M., A.L., B.G., and T.A.P performed the formal analysis. Funding was acquired by B.G. and M.S.H. Investigation was by J.H.H, B.G., and T.A.P. Methodology was developed by J.H.H., M.M., A.L., R.N, B.G, and T.A.P. Project administration was by J.H.H. and T.A.P. J.H.H worked with the software. T.A.P and B.G. supervised the project. J.H.H, B.G., and T.A.P wrote the original draft of the manuscript. All authors contributed to the final version of the manuscript.

## Materials and Correspondence

Please direct all requests to (JHH) and (TAP).

## Notes

### Competing Interest Statement

The authors have declared no competing interest.

