## Supplemental Methods and Results for "Inferring person-to-person networks of *Plasmodium falciparum* transmission: is routine surveillance data up to the task?"

### 1. Methods

#### 1.1. Data

Eswatini is a low-transmission setting in sub-Saharan Africa believed to have high rates of malaria importation[1]. A rigorous surveillance system was established to support the country in making decisions about how to target and tailor interventions for malaria elimination.

Between January 1st, 2013 and December 31st, 2016, 775 cases were identified through active case investigation (ACI, following up on symptomatic malaria cases presenting to health facilities) and reactive case detection (ACD, actively screening neighbors of symptomatic malaria cases) by the National Malaria Elimination Programme (Fig S1A, S1B). ACI were identified using rapid diagnostic test (RDT) in patients presenting with fever to health facilities. Infection status was confirmed with loop-mediated isothermal amplification (LAMP) using a dried blood spot (DBS) which per national policy, was to be collected in all RDT-positives using a second finger prick at the time of presentation and before antimalarial treatment. When DBS was not collected at presentation, a team aimed to visit the patient within 48 hours and collected a DBS at that time. In the analysis, we included all ACIs with a positive LAMP result as well as ACIs with a negative LAMP result if the DBS was collected following treatment, due to the rapid decline of parasitemia after treatment. ACD refers to largely asymptomatic RDT and LAMP-positive individuals identified through reactive case detection (malaria testing using RDT for household members and neighbors of passive detected index cases). We included ACDs with a positive LAMP result as well as ACDs with a positive RDT result and no LAMP result. Two LAMP-positive cases with missing RDT results were excluded from the final dataset.

For the inference algorithm, we assumed that each ACI was symptomatic and received treatment, in keeping with local procedures. We further assumed that each ACD was asymptomatic and received treatment upon identification if a positive RDT result was obtained, in keeping with local procedures. Otherwise, the ACD was assumed to be untreated. In total, 676 cases were symptomatic and treated, 42 cases were asymptomatic and treated, and 57 cases were asymptomatic and untreated (Fig S1C). The timing of detection was recorded for each case, and the spatial coordinates of the location of detection was available for 762 cases and missing for 13 cases. At the time of the case investigation, which generally took place within 48 hours of diagnosis, patients were asked to provide a detailed travel history of all travel outside of their village within the 8 weeks prior to presentation. The travel locations (country, region, town) and dates of travel for up to 5 trips were also collected. Due to the at least one week incubation period needed for *P. falciparum*, only travel in the 1 to 8 weeks prior to presentation was considered as a potential source for the infection. Local versus imported classification was determined by the National Malaria Elimination Programme. If a case classification was not available, we assigned a positive travel history to cases reporting travel outside Eswatini during the 1 to 8 weeks prior to detection. In total, 55% of cases (n=423) had positive travel histories, 41% (n=316) had negative travel histories, and 4% (n=36) had unknown travel histories (Fig S1D).

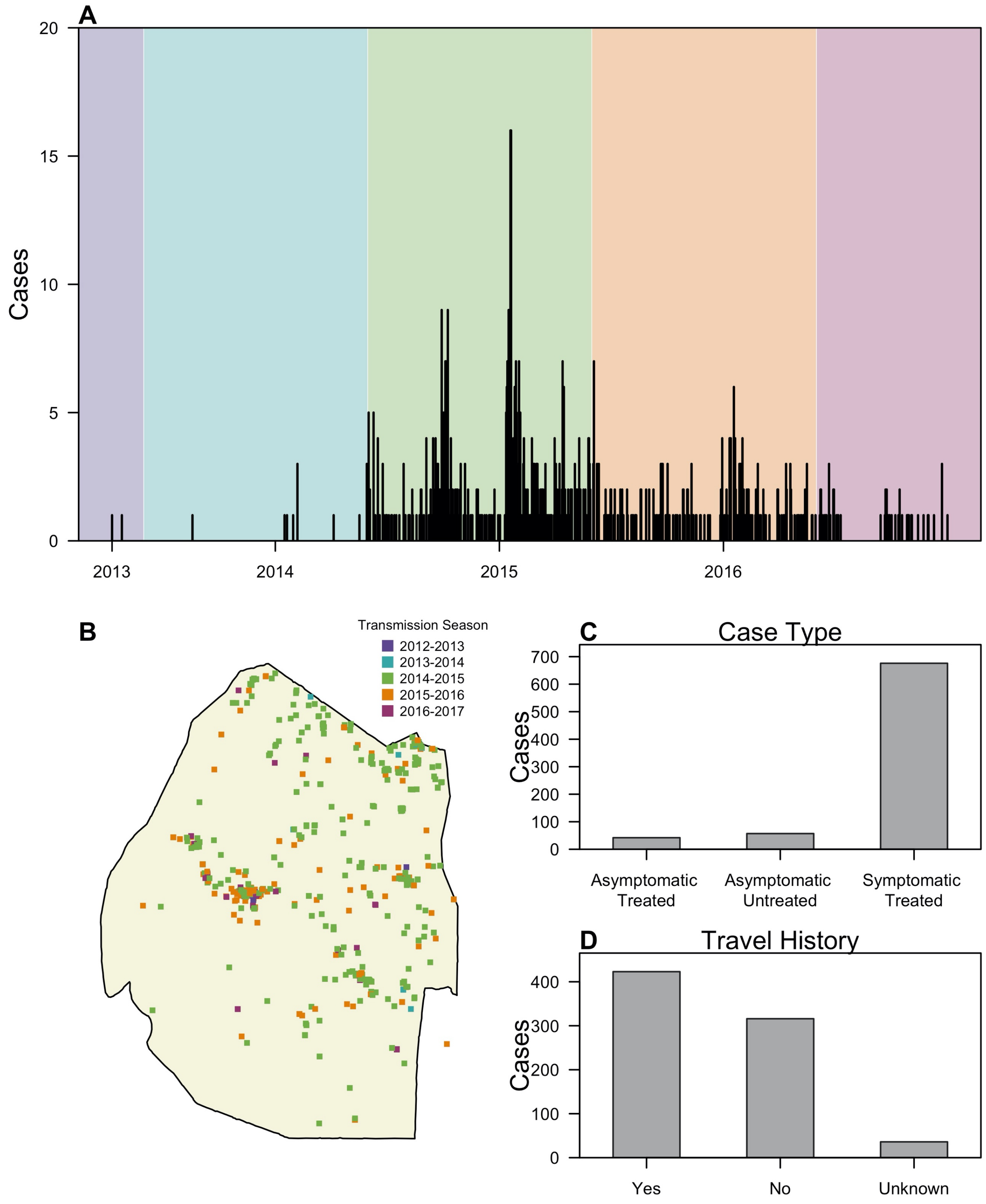

***S1 Fig. Summary of P. falciparum cases in Eswatini.*** *(A) The time series of cases during 2013-2016 is shown with the regions color-coded according to the corresponding transmission season.*

*The transmission season for each year begins on June 1st and ends on May 31st of the following year, and case data was available for the 2012-2013 (purple), 2013-2014 (blue), 2014-2015 (green), 2015-2016 (orange), and 2016-2017 (maroon) transmission seasons. (B) Cases are mapped according to the location of detection and color-coded by the transmission season during which they were detected. (C) The number of cases that were asymptomatic and untreated, asymptomatic and untreated, and symptomatic and treated is reported. (D) The number of cases that a positive travel history (Yes), a negative travel history (No), or unknown travel history (Unknown) is reported.*

#### 1.2. Analysis

##### *1.2.1. Validation using a simple test case*

To demonstrate the validity of our algorithm in a basic sense, we constructed three idealized test cases. Each represented a best-case scenario in which we believed that our algorithm should perform properly if implemented correctly. These involved networks consisting of 20 cases, where the proportion of imported cases was 5%, 50%, or 90%. To further simplify the inference exercise, we simulated under a perfectly accurate travel history (i.e., $\tau_s = 1$ and $\tau_l = 0$). The serial interval between cases involved in each local transmission event was fixed at the mean value of 49 days. Furthermore, the spatial arrangement of cases, both within and between outbreaks, was chosen to represent an ideal transmission network. Specifically, imported cases were distributed according to a Poisson process within a disk with radius of 100 km, ensuring that outbreaks were spatially isolated. Moreover, the spatial distribution of nodes within each outbreak was selected using the Kamada-Kawai algorithm[2] to ensure sufficient spatial separation between cases not linked by transmission. Finally, we assigned the coordinates of each node such that the spatial distribution of the nodes was maintained and the distance separating a transmission event was

2.5 km. This distance is the mean of a half-normal distribution with variance given a serial interval of 49 days and a diffusion coefficient of 0.2 (taken as the geometric mean of the upper and lower bounds of diffusion coefficients considered by Reiner *et al.*[3]).

We performed inference on the three simulated data sets under five different combinations of data types and assumptions about the travel history (S1 Table). These inference settings used: (1) spatial and temporal data while estimating the accuracy of the travel history (default setting); (2) spatial and temporal data while believing the travel history (as in Reiner *et al.*[3] and Routledge *et al.*[4]); (3) spatial and temporal data alone; (4) temporal data while estimating the accuracy of the travel history; and (5) temporal data while believing the travel history (as in Routledge *et al.*[5]). We then measured the accuracies of case classification as imported or locally acquired, identifying a transmission linkage, identifying the correct outbreak of each locally acquired case, and estimating $R_c$.

**S1 Table. Summary of inference settings used.**

| Inference Setting | Spatial Data | Temporal Data | Travel-History Data | | |
|---|---|---|---|---|---|
| | | | Estimate Accuracy | Believe | Ignore |
| 1 (Default) | X | X | X | | |
| 2 | X | X | | X | |
| 3 | X | X | | | X |
| 4 | | X | X | | |
| 5 | | X | | X | |

*The choice of data types and assumptions about the travel history are provided for each of the five inference settings considered. For each setting, an "X" indicates that corresponding data type was included or assumption about the travel history was made.*

*1.2.2. Application to Eswatini surveillance data*

After validating our algorithm using a simple test case, we applied it to malaria surveillance data from Eswatini. These data consist of household location, timing of clinical presentation, the presence or absence of symptoms, treatment status, and self-reported travel histories of 775 cases investigated by the National Malaria Elimination Programme of Eswatini during 2013-2017.

To determine how transmission network inferences depended on the inclusion of various data types and assumptions about travel-history accuracy, we applied our algorithm to the Eswatini surveillance data under the five different inference settings outlined in S1 Table. After assessing convergence, we examined the sensitivity of parameter estimates and the inferred spatial and temporal scales of transmission to these inference settings. Additionally, we considered how estimates of epidemiologically relevant quantities, including $R_c$ and the proportion of cases that were imported, depended on these inference settings. We mapped the latter two quantities across Eswatini using a generalized additive model with a Gaussian process basis function setting using the *mgcv* package in R[6,7]. To ensure that the responses variables were approximately Gaussian-distributed, we took the natural logarithm of the individual-level mean

$R_c$ estimates for each node, padded by $10^{-5}$ to account for zeros. Similarly, we transformed the probability that each case was imported using the approach of Smithson and Verkuilen[8].

*1.2.3. Validation of inferences from Eswatini*

To validate our inferences on Eswatini surveillance data, we applied our algorithm to simulated data generated using the median posterior parameter estimates inferred from the surveillance data and evaluated our ability to recover known networks and parameter values. We did this by simulating under and inferring under the same inference setting, for all five inference settings. The goal of these exercises was to understand the potential limits of the accuracy of our inferences on the Eswatini surveillance data, where the true network and parameters were unknown. As with the simple test case, we measured the accuracy of classifying cases as imported or locally acquired, inferring transmission linkages, identifying the correct outbreak for each locally acquired case, and estimating $R_c$.

For this exercise, we simulated transmission networks and corresponding epidemiological data (e.g., household location, timing of clinical presentation, etc.) using a branching process model for which generative processes for spatial, temporal, and travel-history data mirrored the assumptions used in the formulation of our likelihood. To simulate data using the branching process, the maximum number of cases (i.e., treated *P. falciparum* infections) and $R_c$ were first specified. We then calculated the maximum number of infections based on the probability of treatment given symptoms (1.0), the probability of treatment given no symptoms (0.42), and the probability of symptoms (0.87). Each probability was calculated empirically from the Eswatini data set. The number of imported infections was equal to the product of the maximum number of infections and the importation proportion (i.e., $1 - R_c$). We uniformly distributed the imported infections over a temporal window consistent with that of the Eswatini data set (1361 days), and we randomly sampled the spatial coordinates of these imported infections according to population density estimates from WorldPop[9]. While the number of treated *P. falciparum* infections was less than the specified maximum number of cases, we sampled the number of offspring from each node according to a Poisson distribution with a mean of $R_c$. For each offspring, the timing and location of detection were sampled relative to the timing and location of detection of the parent using the spatial and temporal kernels formulated in the likelihood. Travel histories for imported and locally acquired cases were Bernoulli trials with probabilities of $\tau_s$ and $\tau_l$, respectively, the symptom status of each case was a Bernoulli trial with the probability of symptoms, and the treatment status of each was a Bernoulli trial with either the probability of treatment given symptoms or the probability of treatment given no symptoms.

Each simulated data set was generated to approximate characteristics of the Eswatini surveillance data along with inferred parameters from the model. Specifically, the number of nodes in the simulated data approximated the total number of cases in the Eswatini surveillance data, and we set the proportion of imported cases ($p_i$), the diffusion coefficient ($D$), and the parameters that govern the accuracies of the travel history ($\tau_s$ and $\tau_l$) to their median values from the posterior distribution. Similarly, under inference settings where the accuracy of the travel history was not estimated, we assigned $\tau_s = 1$ and $\tau_l = 0$, implying perfectly accurate travel histories. To match the observation from surveillance data that individuals who reported travel tended to be located in metropolitan areas, we distributed the imported cases spatially proportional to gridded population density estimates from WorldPop[10].

###### *1.2.4. Simulation Sweep*

To identify the epidemiological parameters that affect the accuracy of reconstructing transmission networks using routinely collected surveillance data, we performed a simulation sweep in which we varied the following epidemiological parameters: (1) the diffusion coefficient, (2) the proportion of imported infections, (3) the temporal window over which imported infections were distributed, (4) the degree of spatial clustering among imported infections[11], (5) $\tau_s$, and (6) $\tau_l$. We sampled 2,000 values for each epidemiological parameter using a Sobol design (S2 Table)[12]. We then parameterized a branching process model with each parameter set to generate a total of 2,000 simulated data sets, each comprising a transmission network of 200 nodes. The number of nodes in each simulated data set was less than the number of nodes in the Eswatini surveillance data and was selected to reduce computational burden. Nevertheless, the relative epidemiological features of the transmission network should affect the accuracy of network reconstruction more so than the size of the network itself. Therefore, we expect that the results of this simulation sweep should generalize to networks of various sizes.

**S2 Table. Parameter ranges for the simulation sweep.**

| Parameter | Range |
|---|---|
| Diffusion Coefficient | (0,30] |
| Proportion of cases that are imported | (0,1] |
| Maximum date of an imported case (days) | (0, 9125] |
| Degree of clustering of imported cases | (0,1] |
| $\tau_s$ | [0,1] |
| $\tau_l$ | [0,1] |

*The parameters and their respective ranges are reported. A Sobol design was used to sample 2,000 parameter values from the respective range and generate simulated data sets*[12].

We applied our inference algorithm under three inference settings to each simulated data set and measured the accuracy of reconstructing transmission networks. The three inference settings used: (1) spatial and temporal data while estimating the accuracy of the travel history (default setting); (2) spatial and temporal data while believing the travel history; and (3) spatial and temporal data alone (S1 Table). We chose to use those inference settings, because they included each of the three assumptions about travel-history data. As with previous validation exercises, we measured the accuracy of classifying cases as imported or locally acquired, inferring transmission linkages, identifying the correct outbreak of each locally acquired case, and estimating $R_c$. We then examined how each of these accuracy metrics varied as a function of the epidemiological parameters.

### 2. Results

#### 2.1. Validation using a simple test case

We first validated our approach on three small, simulated networks of twenty nodes. Although these networks varied in their proportion of imported cases, the local transmission chains were arranged to ensure that there was sufficient spatiotemporal separation between transmission chains, and we simulated perfect travel histories and complete observation of cases, providing idealized test cases to validate our inference algorithm. We measured the performance of our inference algorithm in terms of its ability to reconstruct different features of the transmission network and correctly estimate $R_c$. As the proportion of imported cases decreased from 85% to 5%, we found that the ability of the algorithm to correctly classify cases as imported or locally acquired improved (Fig S2). For example, when we used the default inference setting, classification accuracy improved from 85.7% (95% Credible Interval: 85.7 – 85.7%) to 100% (100 – 100%). As classification accuracy improved, our estimates of $R_c$ also improved, with all five inference settings yielding accurate estimates when imported cases comprised only 5% of total cases (Fig S2C). Similarly, the ability to identify the correct parent of each locally acquired case and assign it to the correct outbreak depended on the extent of local transmission in the network. When 85% of the cases were imported, performance was variable across inference settings (Fig S2A). Using spatial data and either estimating or ignoring the travel histories, the algorithm identified a local optimum in the likelihood and consequently classified all locally acquired cases as imported, leading to highly inaccurate transmission network inferences. Believing the travel history, regardless of whether spatial data was included, enabled us to perfectly reconstruct the transmission network, because the travel-history data was simulated to be perfectly accurate, allowing for correct classification of cases as imported or locally acquired. However, as the proportion of imported cases decreased, the benefit of believing the travel history diminished, and using all data types resulted in the greatest accuracy. For example, in the most extreme case in which only 5% of cases were imported, the accuracy of identifying the true parent ranged from 84.2% (68.4 – 94.7%) under the default inference settings to 63.2% (47.4 – 78.9%) using temporal data and estimating the accuracy of the travel history (Fig S2C). In terms of identifying the outbreak to which a case belongs, the algorithm was accurate under all inference settings, since there was only one outbreak (Fig S2C).

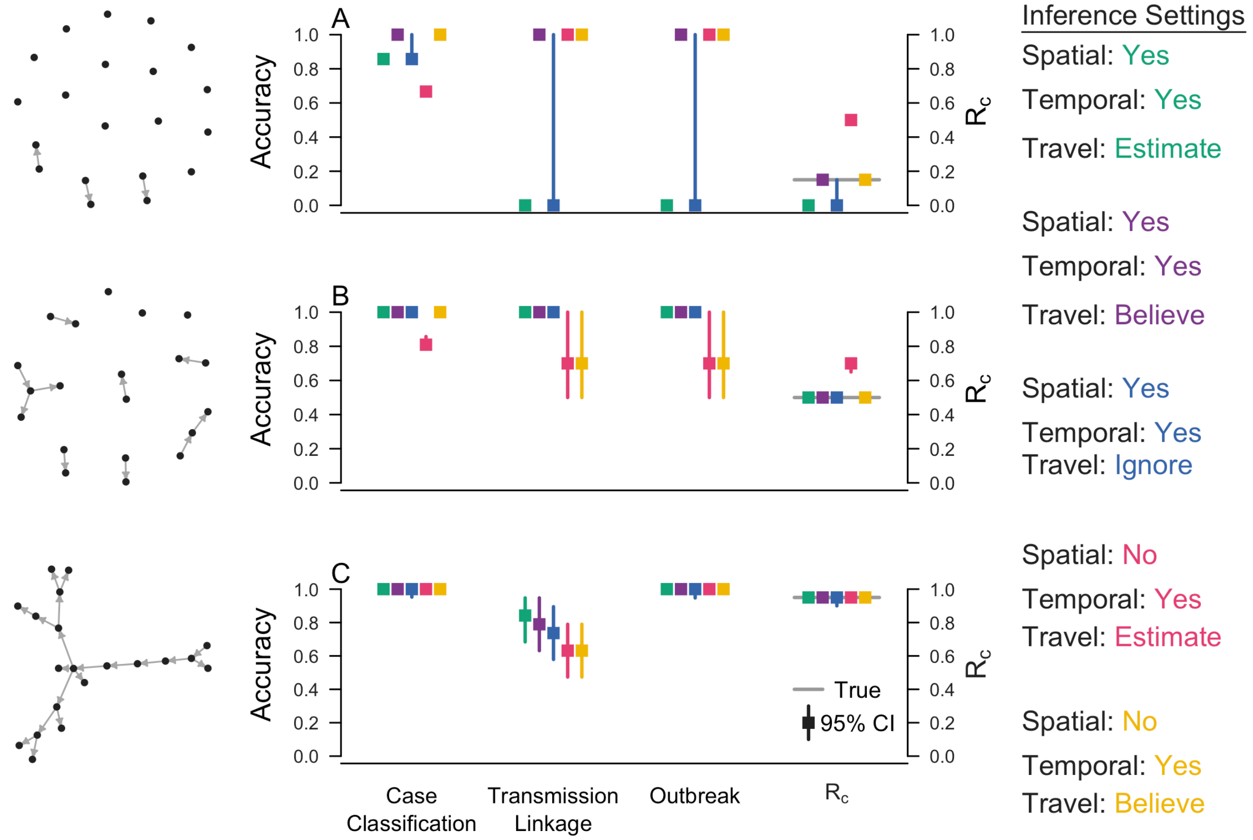

***S2 Fig. Inference accuracies for three simulated transmission networks with (A) 85%, (B) 50%, and (C) 5% of cases as imported.*** *Case classification refers to proportion of cases that are correctly classified as imported vs. locally acquired. Transmission linkage denotes the proportion of locally acquired cases for which the true parent is correctly identified, Outbreak is the proportion of locally acquired cases for which the inferred parent belongs to the correct outbreak, and* $R_c$ *is the estimated reproduction number under control. Square points signify the median posterior value, and bars are the 95% credible intervals. The gray line indicates the true value of* $R_c$*.*

#### 2.2. Likelihood Profile of the Diffusion Coefficient

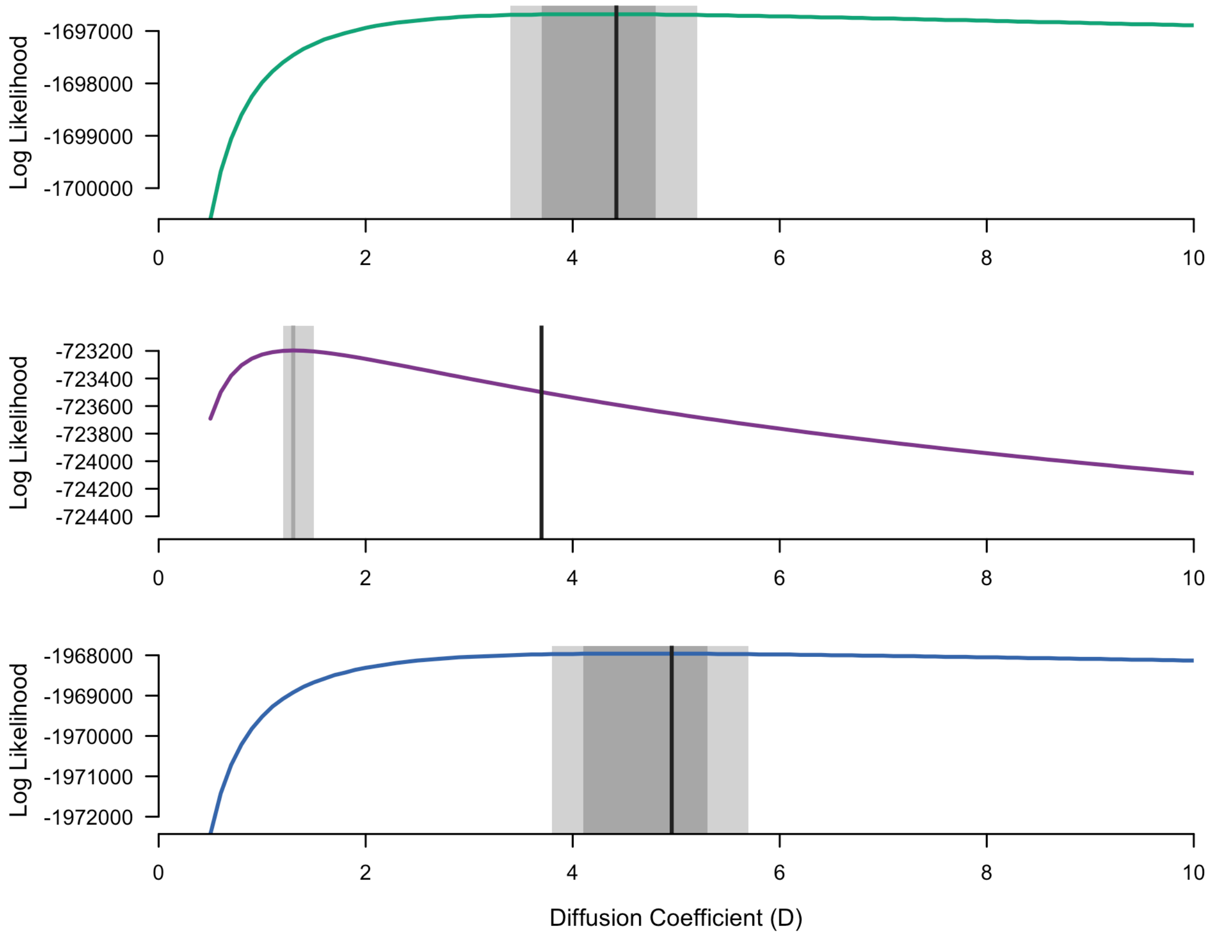

***S3 Fig. Likelihood profile of the diffusion coefficient conditioning on the true network.*** *The likelihood profile for two inference settings that incorporate spatial data are shown as a function of the diffusion coefficient. The green line corresponds to the default inference setting, the purple line corresponds to the setting in which spatial and temporal data are used and the travel history is believed, and the blue line corresponds to the setting in which spatial and temporal data are used and the travel history was ignored. Black bars denote the true value of the diffusion coefficient, dark grey shapes denote the region with the maximum likelihood, and light grey shapes denote the region within 10 log likelihood units of the maximum likelihood.*

#### 2.3. Convergence of Posterior Transmission Network Inferences

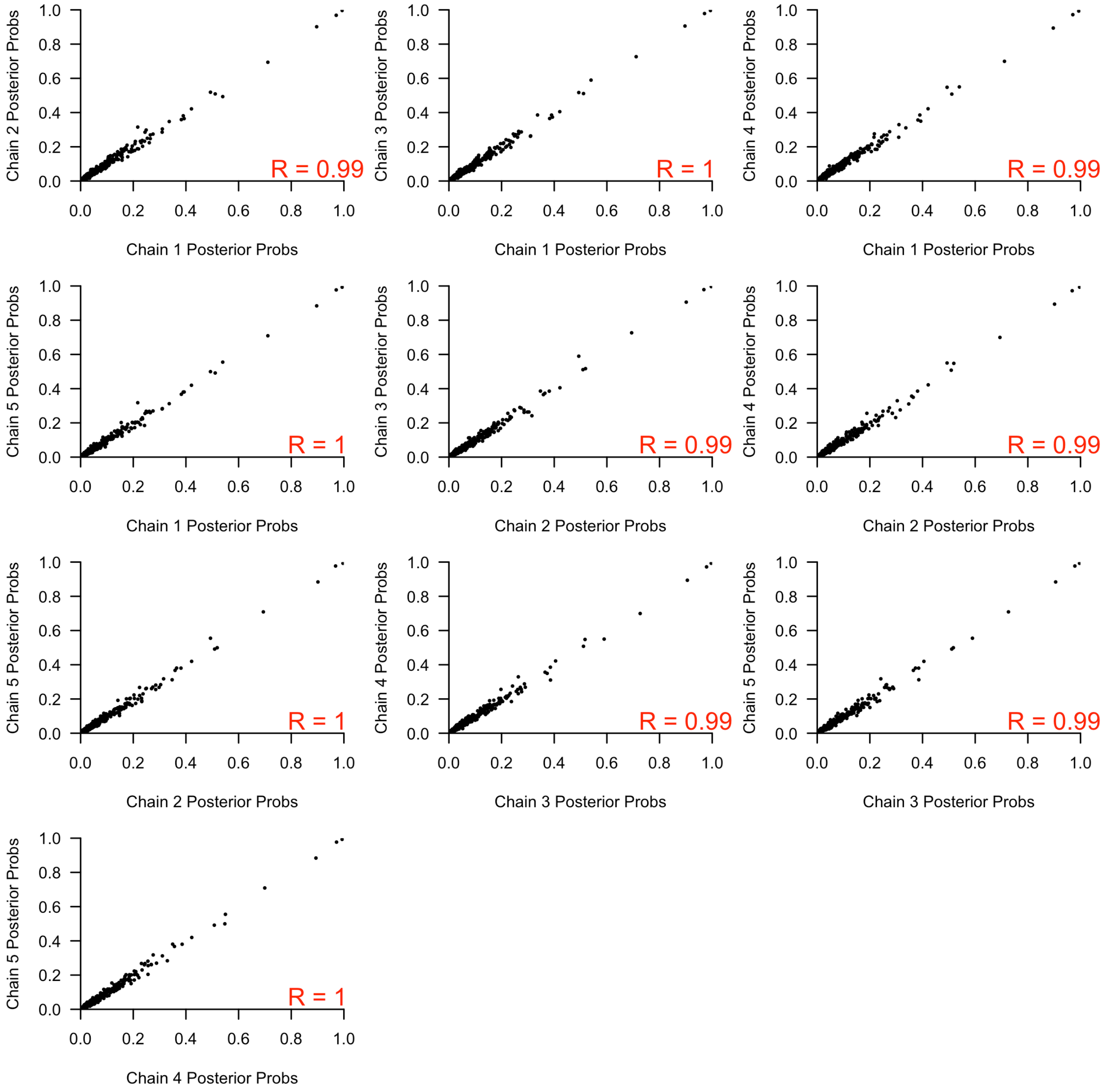

***S4 Fig. Comparison of individual-level importation probabilities on Eswatini data under default settings.*** *Pairwise scatter plots of importation probabilities are shown for each pairing of the 5 independent replicates to assess convergence. The correlation between importation probabilities for each pair of replicates is reported in red.*

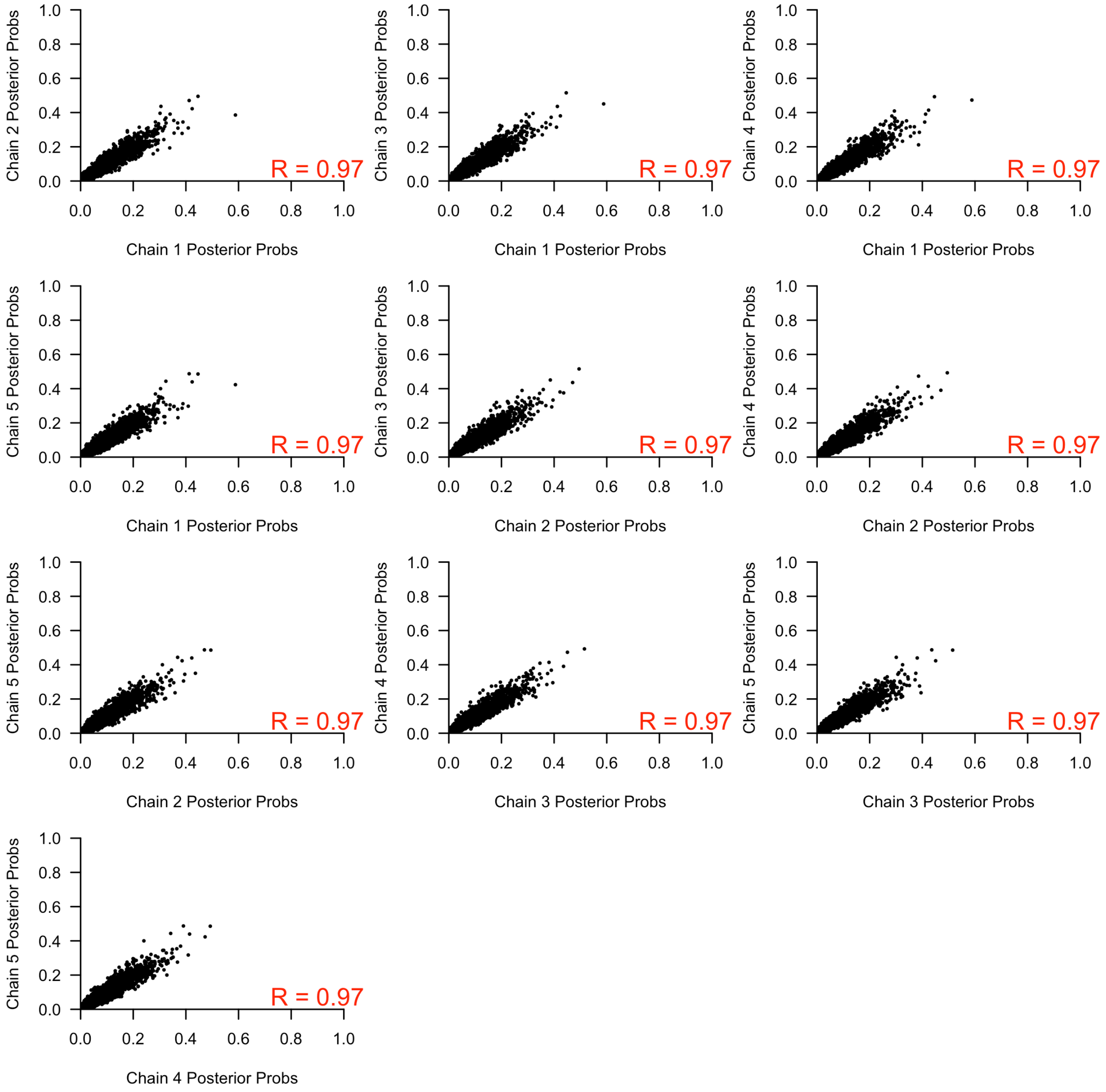

***S5 Fig. Comparison of transmission linkage probabilities on Eswatini data under default settings.*** *Pairwise scatter plots of the probability of each transmission linkage are shown for each pairing of the 5 independent replicates to assess convergence. The correlation between transmission linkage probabilities for each pair of replicates is reported in red.*

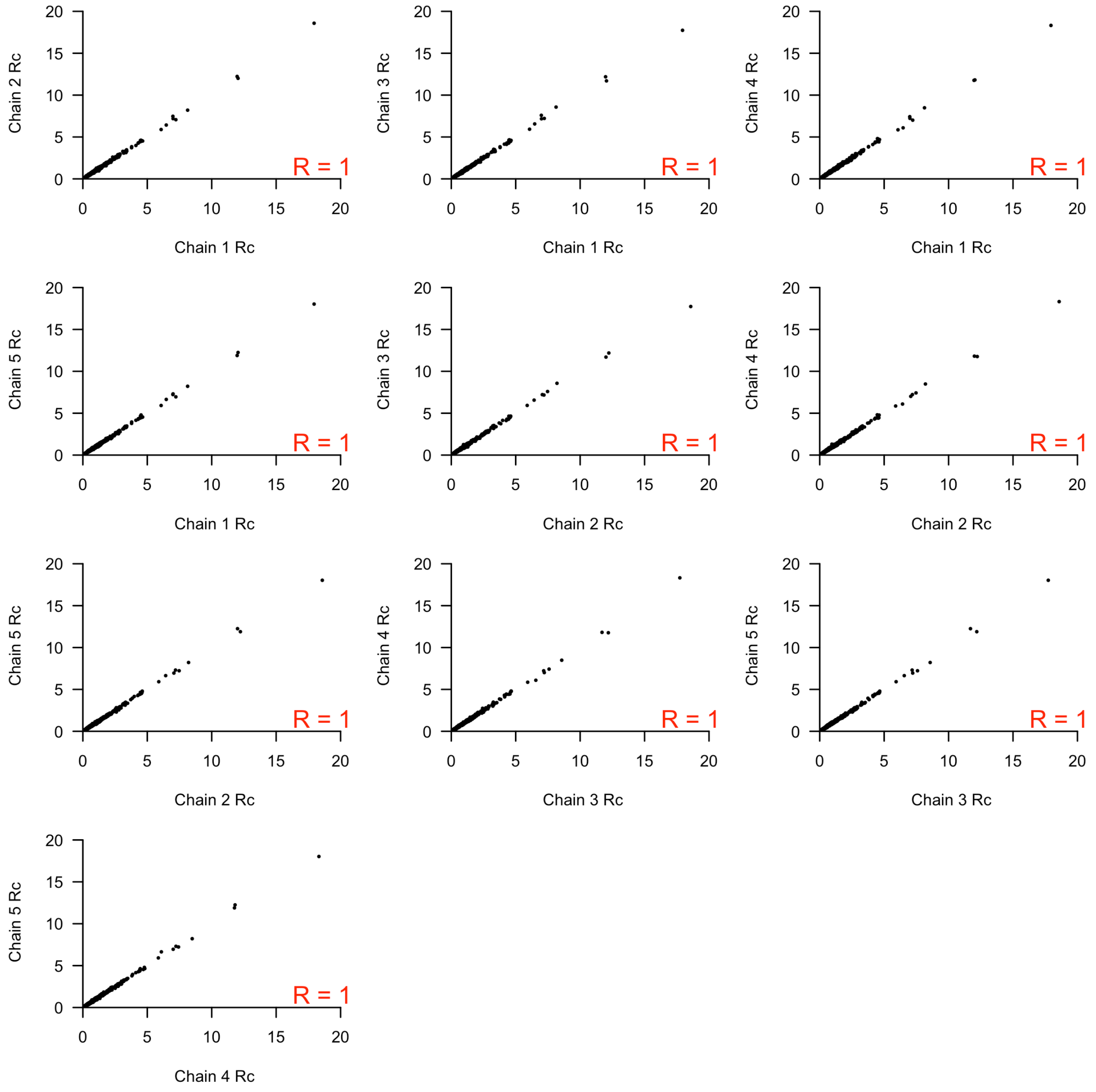

***S6 Fig. Comparison of individual-level $R_c$ estimates on Eswatini data under default settings.*** *Pairwise scatter plots of individual-level $R_c$ are show for each pairing of the five independent replicates to assess convergence. The correlation between individual-level $R_c$ values is reported in red.*

#### 2.4. Analytical Solution for the Posterior Distribution of $\tau_s$ and $\tau_l$

We specified a Bernoulli likelihood and a beta-distributed prior on $\tau_s$ and $\tau_l$. Therefore, the posterior distributions of $\tau_s$ and $\tau_l$ satisfy a conjugate-prior relationship and can be solved analytically as beta distributions.

The prior distribution for $\tau_s$ is a beta distribution with hyperparameters $\alpha_s = 12$ and $\beta_s = 3$, and the prior distribution for $\tau_l$ is a beta distribution with hyperparameters $\alpha_l = 3$ and $\beta_l = 12$. Following the conjugate-prior relationship, the posterior distribution for $\tau_s$ is calculated as a beta distribution with hyperparameters,

$$\hat{\alpha}_s = \alpha_s + \sum_{n_{founders}} \mathbb{I}(report\ travel) \quad (S1)$$

$$\hat{\beta}_s = \beta_s + \left( n_{founders} - \sum_{n_{founders}} \mathbb{I}(report\ travel) \right). \quad (S2)$$

The summation in eq. (S1) is the number of cases that reported travel and are inferred by the algorithm to be imported cases. Similarly, the second term in eq. (S2) is the number of cases inferred by the algorithm to be imported cases that did not report travel. The posterior distribution for $\tau_l$ is similarly described by a beta distribution with hyperparameters,

$$\hat{\alpha}_l = \alpha_l + \sum_{n_{local}} \mathbb{I}(report\ travel) \quad (S3)$$

$$\hat{\beta}_l = \beta_l + \left( n_{local} - \sum_{n_{local}} \mathbb{I}(report\ travel) \right). \quad (S4)$$

In eq. (S3), the summation is the number of cases that reported travel and were inferred by the algorithm to be locally acquired. Similarly, the second term in eq. (S4) is the number of cases inferred by the algorithm to be locally acquired that did not report travel.

We then compared the prior distributions, the posterior distributions obtained from the MC3 sampling algorithm, and the posterior distribution obtained using the analytical solutions in eqs. (S1-S4) for $\tau_s$ and $\tau_l$ inferred from the Eswatini surveillance data. Because each case had a posterior probability of being imported or locally acquired but eqs. (S1-S4) required a binary classification, we classified a case as imported if the posterior probability of being imported exceeded 0.25. This threshold was arbitrarily defined, but the purpose of this exercise is purely illustrative.

Under both inference settings in which the accuracy of the travel histories was inferred, we observed good agreement between the analytical and numerical posterior distributions for $\tau_s$ and $\tau_l$. Whether or not the posterior distribution deviated from the prior distribution depended upon the number of cases that were classified as imported or locally acquired. When there are more cases classified as imported, the strength of the data predominated in eqs. (S1-S2), and the posterior distribution of $\tau_s$ deviated from the prior distribution. By contrast, when most cases are locally acquired, the posterior distribution of $\tau_s$ resembled the prior distribution. This is consistent with the posterior distributions that we observed when we used spatial and temporal data and estimated the accuracy of the travel history versus when we used temporal data and estimated the accuracy of the travel history. Using the former, we estimated 5.2% of the cases as imported, which was sufficient to shift the posterior distribution of $\tau_s$ away from the prior distribution (S7 Fig). Using the latter, we only estimated 0.13% of cases as imported. This small number of imported cases implied that the posterior distribution of $\tau_s$ resembled the prior distribution (S8 Fig).

The derivation of the analytical solution of $\tau_s$ explains our inability to correctly estimate this parameter from simulated data (Fig 4B). Using the spatial and temporal data and estimating the accuracy of the travel history, the true value of $\tau_s$ was 0.61, and 5.2% of all cases in the simulated data set were imported. However, applying the MC3 algorithm to this simulated data set, we inferred only ~1% of all cases to be imported. Consequently, we do not estimate a sufficient number of imported cases to shift the posterior distribution of $\tau_s$ away from the prior distribution and correctly estimate this parameter (S9 Fig).

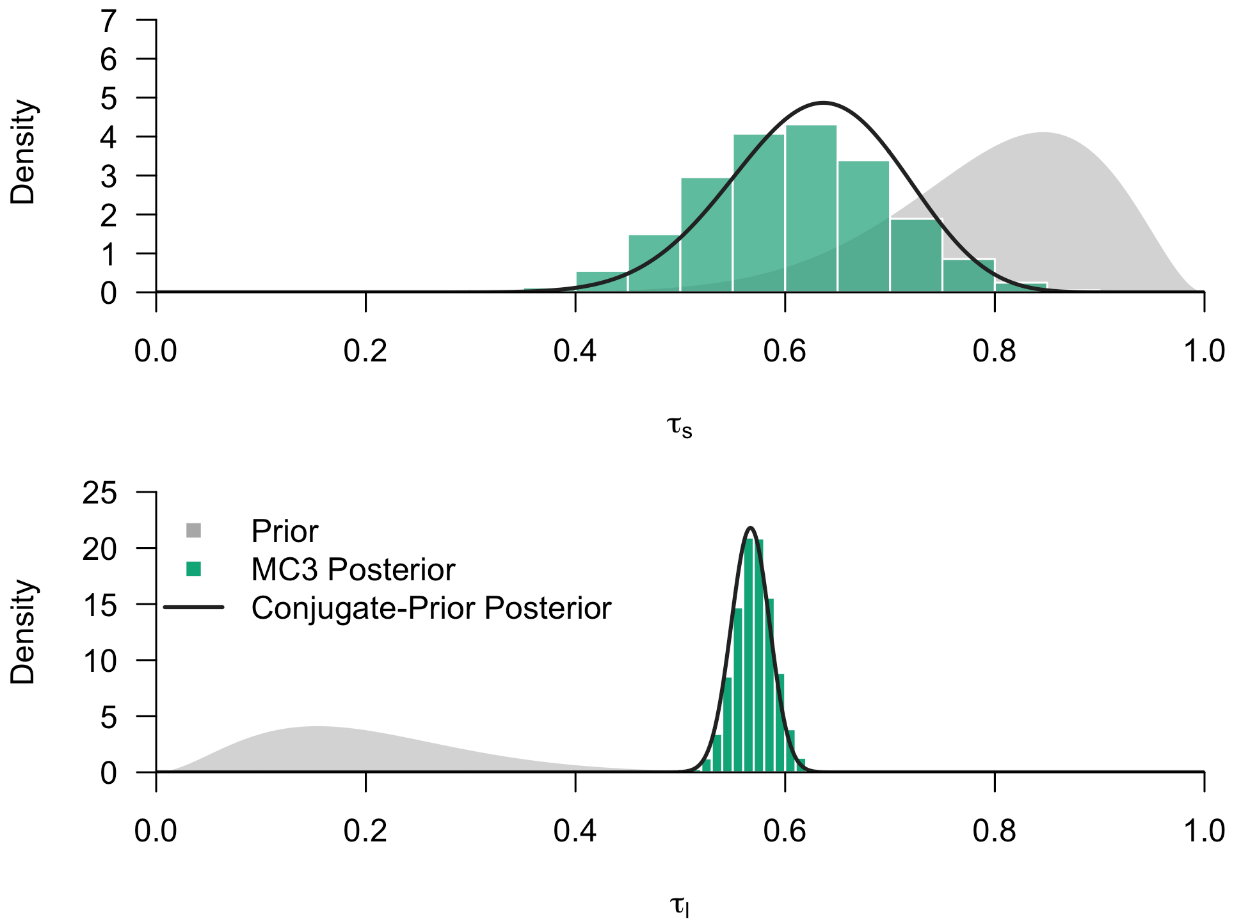

***S7 Fig. Comparison of the prior and posteriors of $\tau_s$ and $\tau_l$ from the Eswatini surveillance data using spatial and temporal data and estimating the accuracy of the travel history.*** *The prior (gray shape), the analytical posterior distribution (black line), and the numerical posterior distribution from MC3 (green histogram) are plotted for $\tau_s$ and $\tau_l$.*

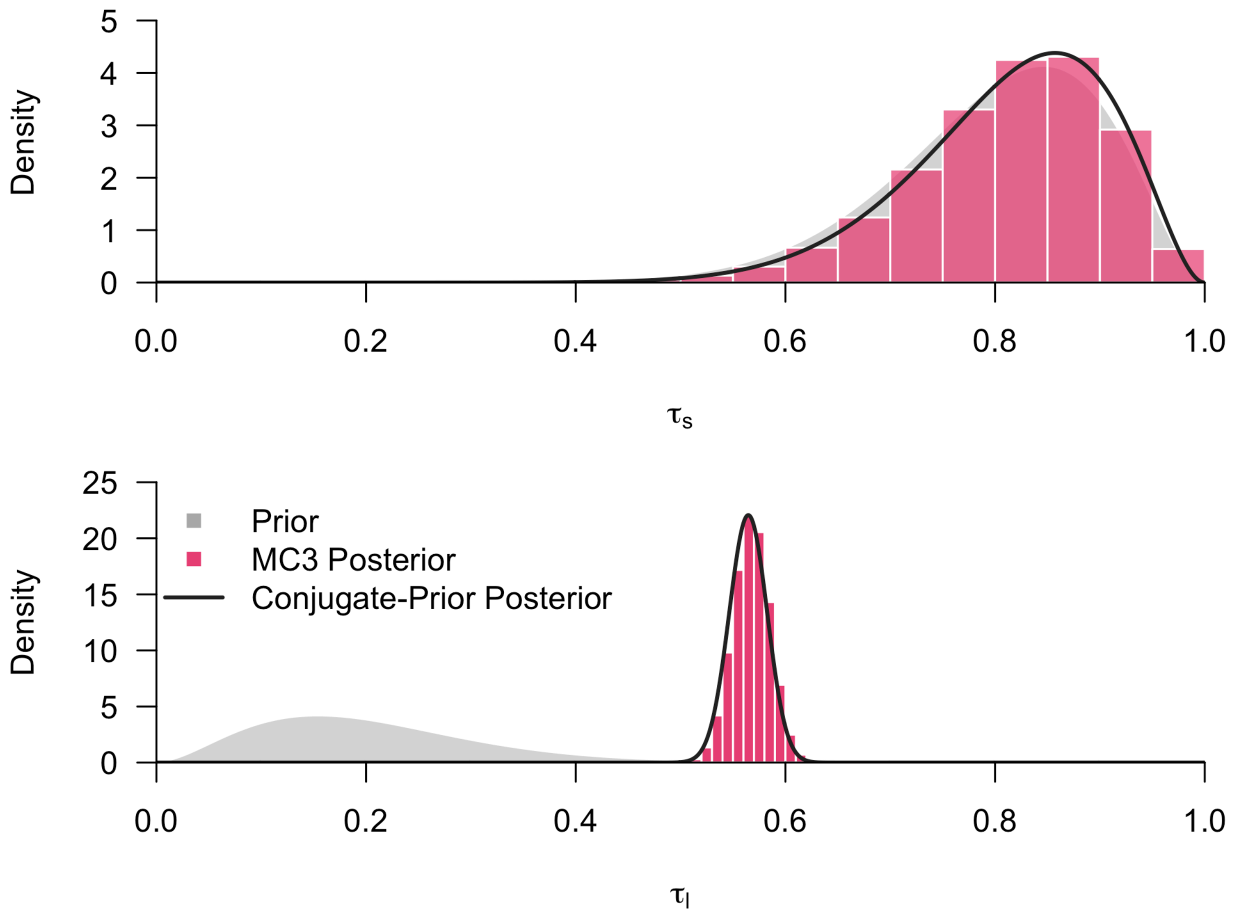

***S8 Fig. Comparison of the prior and posteriors of $\tau_s$ and $\tau_l$ from the Eswatini surveillance data using temporal data and estimating the accuracy of the travel history.*** *The prior (gray shape), the analytical posterior distribution (black line), and the numerical posterior distribution from MC3 (pink histogram) are plotted for $\tau_s$ and $\tau_l$.*

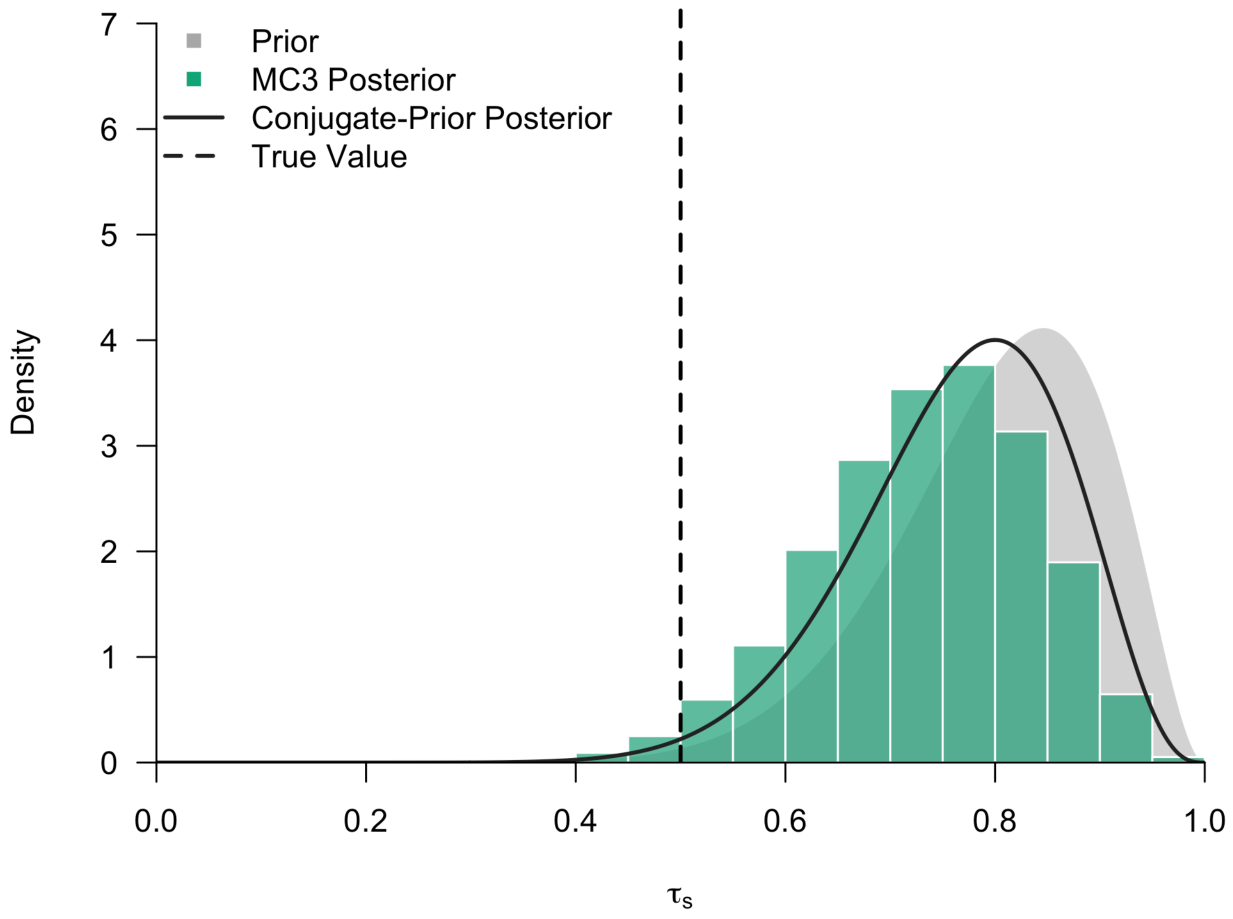

***S9 Fig. Comparison of the prior and posteriors of $\tau_s$ from simulated data using spatial and temporal data and estimating the accuracy of the travel history.*** *The prior (gray shape), the analytical posterior distribution (black line), and the numerical posterior distribution from MC3 (green histogram) are plotted for $\tau_s$.*

#### 2.5. Simulation Sweep

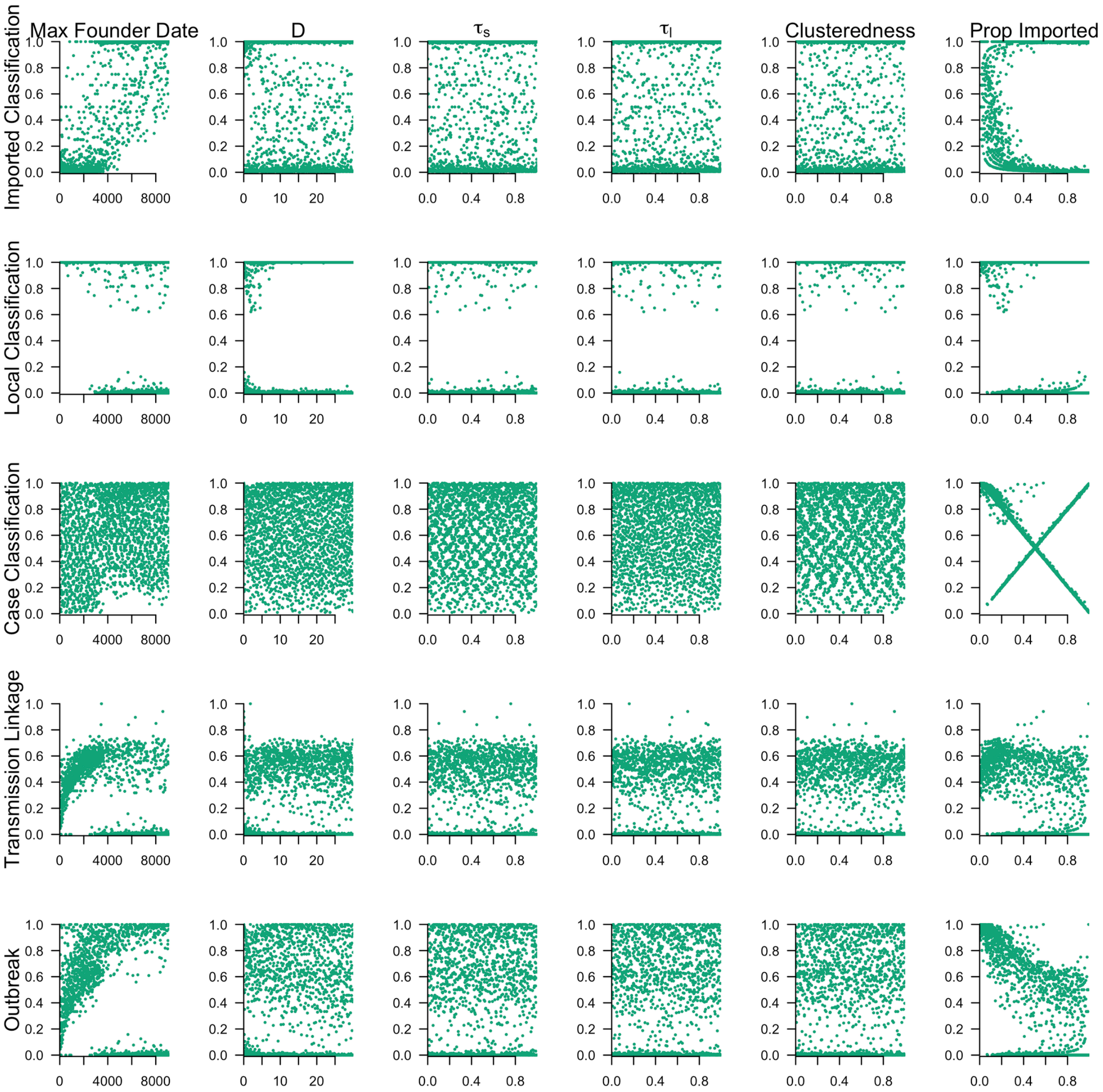

***S10 Fig. Univariate relationships between accuracy metrics and simulation parameters using spatial and temporal data and estimating the accuracy of the travel history.*** *Scatterplots of the relationship between five accuracy metrics and the simulation parameters are reported. Imported classification refers to the proportion of imported cases that are correct classified as imported, local classification refers to the proportion of locally acquired cases that are correctly classified as locally acquired, and case classification refers to the proportion of all cases that*

*are correctly classified as imported or locally acquired. Transmission linkage is the proportion of locally acquired cases for which the true parent is correctly identified, and outbreak is the proportion of locally acquired cases for which the inferred parent belongs to the same outbreak.*

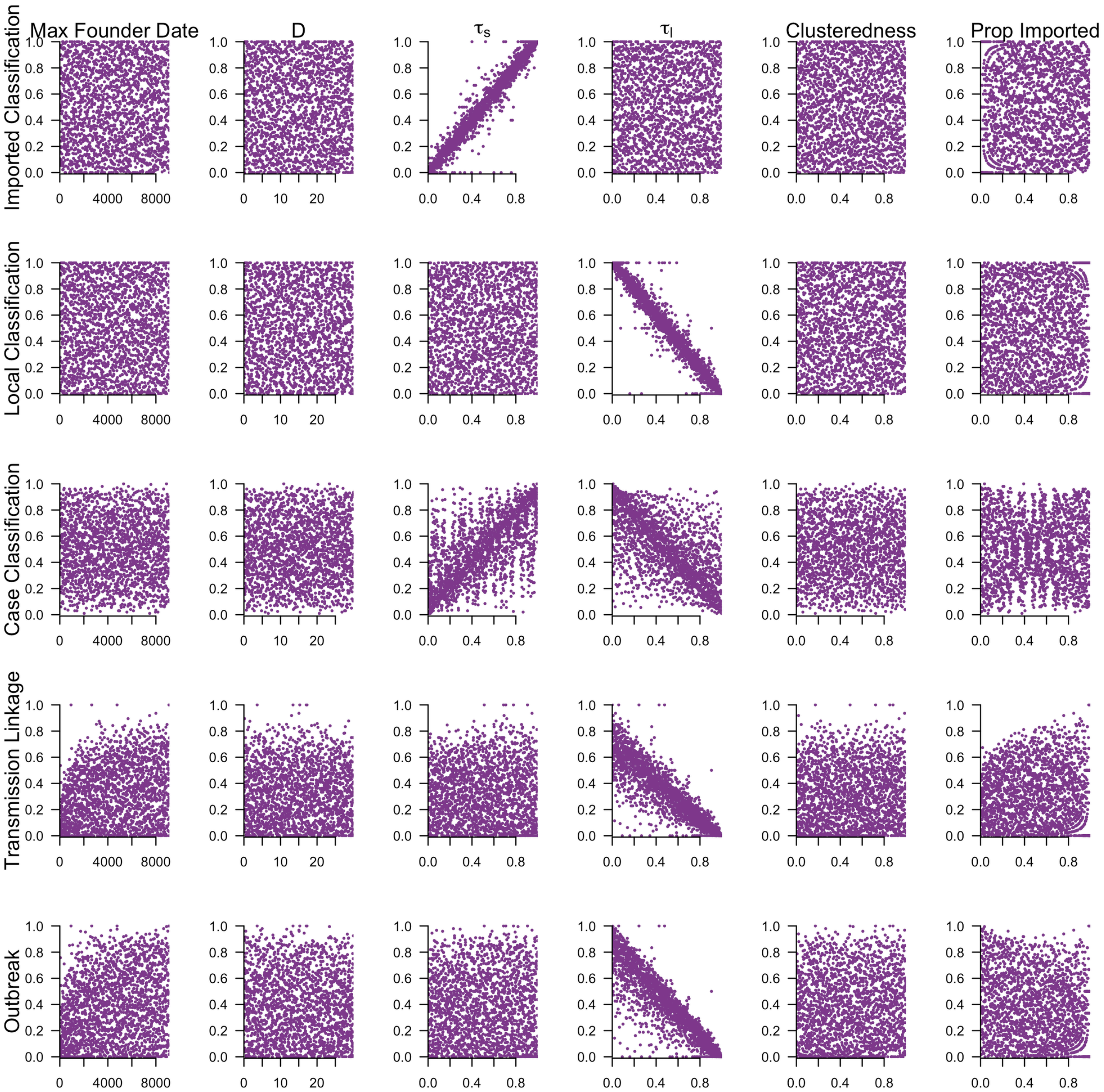

***S11 Fig. Univariate relationships between accuracy metrics and simulation parameters using spatial and temporal data and believing the travel history.*** *Scatterplots of the relationship between five accuracy metrics and the simulation parameters are reported. Imported classification refers to the proportion of imported cases that are correct classified as imported, local classification refers to the proportion of locally acquired cases that are correctly classified as locally acquired, and case classification refers to the proportion of all cases that are correctly*

*classified as imported or locally acquired. Transmission linkage is the proportion of locally acquired cases for which the true parent is correctly identified, and outbreak is the proportion of locally acquired cases for which the inferred parent belongs to the same outbreak.*

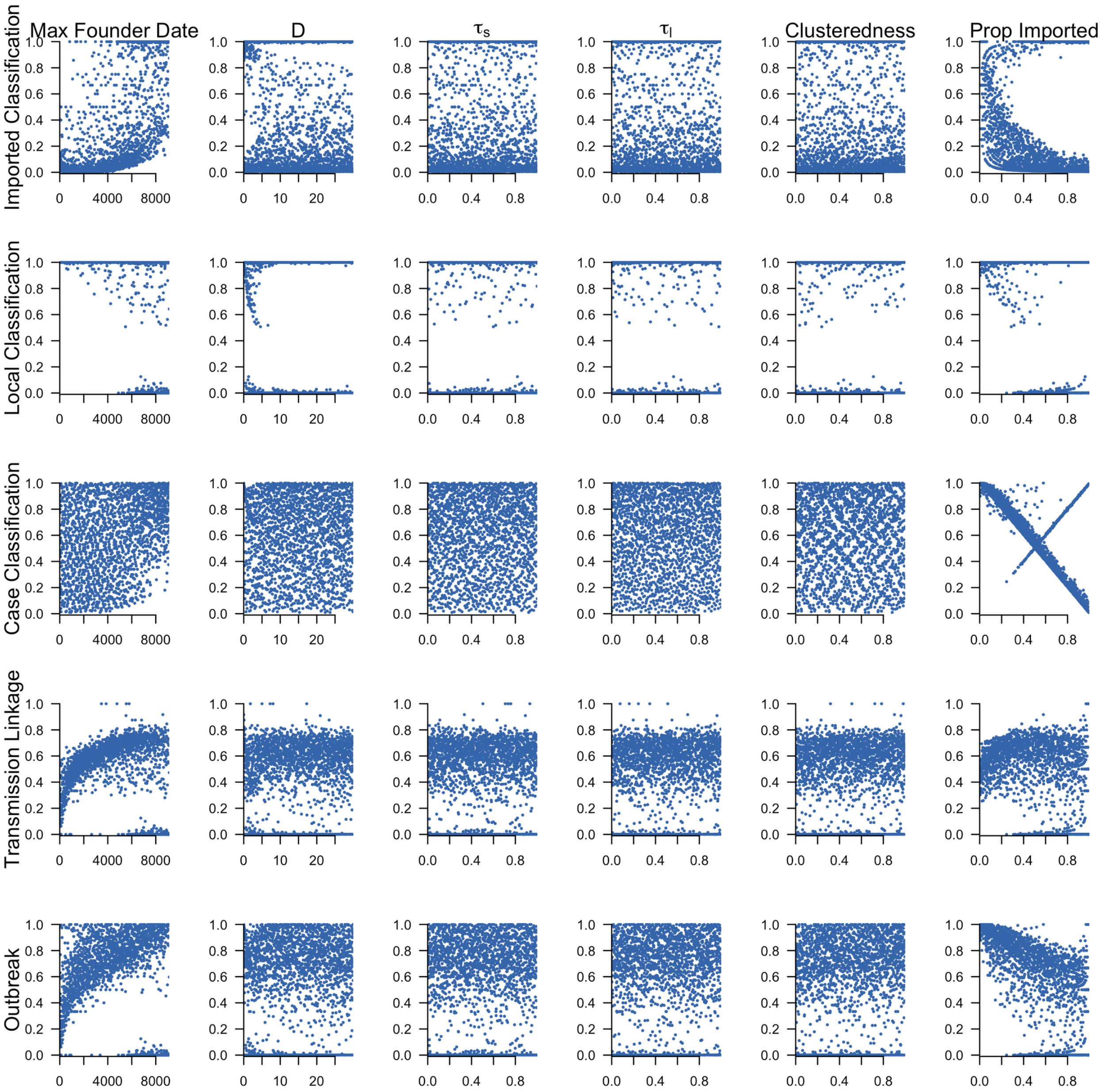

***S12 Fig. Univariate relationships between accuracy metrics and simulation parameters using spatial and temporal data only.*** *Scatterplots of the relationship between five accuracy metrics and the simulation parameters are reported. Imported classification refers to the proportion of imported cases that are correct classified as imported, local classification refers to the proportion of locally acquired cases that are correctly classified as locally acquired, and case classification refers to the proportion of all cases that are correctly classified as imported or*

*locally acquired. Transmission linkage is the proportion of locally acquired cases for which the true parent is correctly identified, and outbreak is the proportion of locally acquired cases for which the inferred parent belongs to the same outbreak.*

#### 2.6. Uncertainty in Higher-Order Summaries of the Network

The credible intervals for higher-order summaries of the network, including case classification and $R_c$, are narrow, because the calculation of these higher-order summaries requires that we apply a binary condition to each node in the network (e.g., "Was the node inferred to be imported or locally acquired?", "Was the inferred parent the true parent?", etc.) In doing so, we compressed or reduced much of the uncertainty inherent to the inferred network. Because the estimates of $R_c$ depend only upon the case classification, we obtained narrow credible intervals for $R_c$.

To consider this further, we took the posterior distributions from the "Validation of Inferences from Eswatini" analysis and compared the log-likelihoods that each case was imported or locally acquired. For each of the three inference settings examined (S13-S15 Figs), the log-likelihood that each case was locally acquired was generally higher, because, in each simulated network, there were 775 cases over 1361 days. This ensured that there was generally a plausible observed parent that occurred within one serial interval prior to each case. The log-likelihood that a case was imported was higher for asymptomatic cases than symptomatic cases, because the serial interval distribution was more diffuse for asymptomatic cases[13]. Although this effect is sensitive to our assumption about the different serial interval distribution for symptomatic and asymptomatic cases, the calculation of the serial interval distributions was informed by empirical data collected in Zanzibar and modeled asexual parasite densities obtained from a validated, within-host model of *P. falciparum infection.*

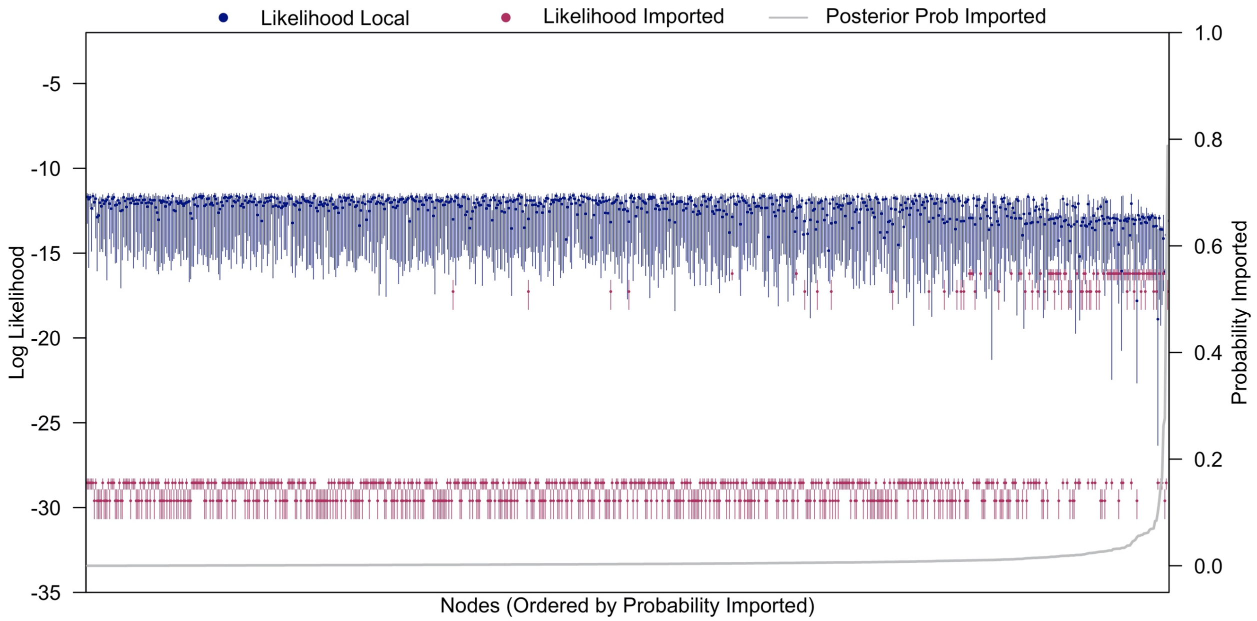

***S13 Fig. Likelihoods of case classification simulated nodes using spatial and temporal data and estimating the accuracy of the travel history.*** *The log likelihoods that each node is locally acquired (navy) or imported (maroon) is calculated for each network from the posterior distribution. Points are the median estimate across the full posterior distribution, and segments are the 95% credible intervals. The gray line is the posterior probability that each node was imported, and the nodes are ordered by increasing posterior probability of importation.*

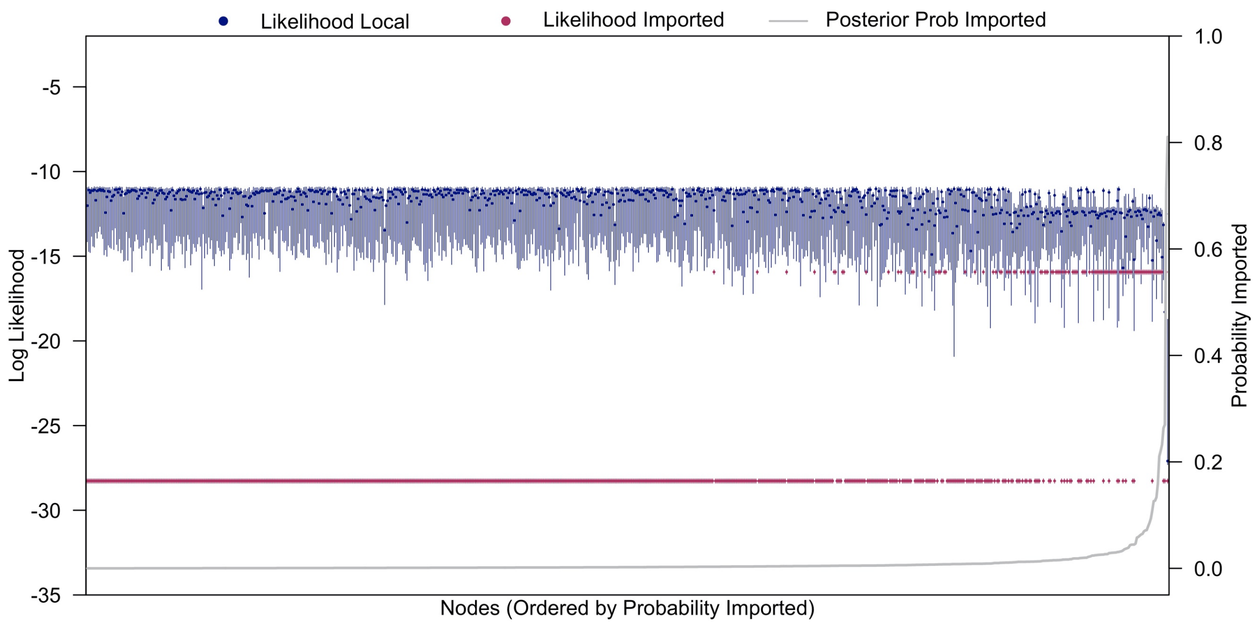

***S14 Fig. Likelihoods of case classification simulated nodes using spatial and temporal data only.*** *The log likelihoods that each node is locally acquired (navy) or imported (maroon) is calculated for each network from the posterior distribution. Points are the median estimate across the full posterior distribution, and segments are the 95% credible intervals. The gray line is the posterior probability that each node was imported, and the nodes are ordered by increasing posterior probability of importation.*

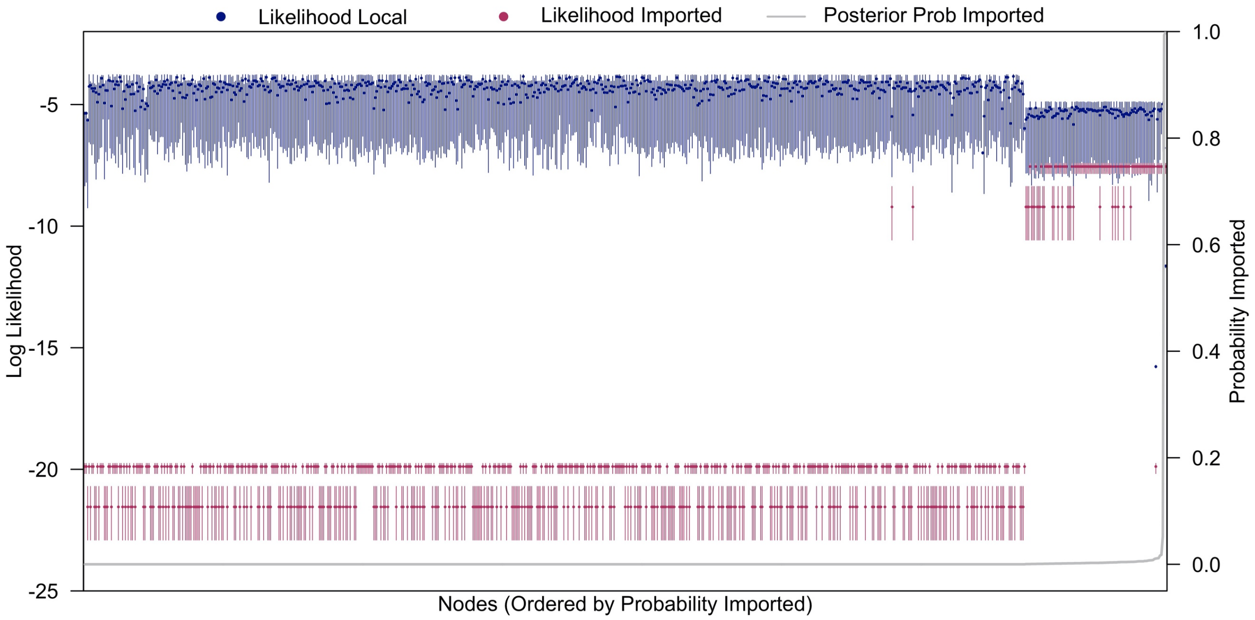

***S13 Fig. Likelihoods of case classification simulated nodes using temporal data and estimating the accuracy of the travel history.*** *The log likelihoods that each node is locally acquired (navy) or imported (maroon) is calculated for each network from the posterior distribution. Points are the median estimate across the full posterior distribution, and segments are the 95% credible intervals. The gray line is the posterior probability that each node was imported, and the nodes are ordered by increasing posterior probability of importation.*
